# HIV-1 latency reversal agent boosting is not limited by opioid use

**DOI:** 10.1101/2023.05.26.23290576

**Authors:** Tyler Lilie, Jennifer Bouzy, Archana Asundi, Jessica Taylor, Samantha Roche, Alex Olson, Kendyll Coxen, Heather Corry, Hannah Jordan, Kiera Clayton, Nina Lin, Athe Tsibris

## Abstract

The opioid epidemic may impact the HIV-1 reservoir and its reversal from latency in virally suppressed people with HIV (PWH). We studied forty-seven PWH and observed that lowering the concentration of HIV-1 latency reversal agents (LRA), used in combination with small molecules that do not reverse latency, synergistically increases the magnitude of HIV-1 re-activation *ex vivo*, regardless of opioid use. This LRA boosting, which combines a Smac mimetic or low-dose protein kinase C agonist with histone deacetylase inhibitors, can generate significantly more unspliced HIV-1 transcription than phorbol 12-myristate 13-acetate (PMA) with ionomycin (PMAi), the maximal known HIV-1 reactivator. LRA boosting associated with greater histone acetylation in CD4^+^ T cells and modulated T cell activation-induced markers and intracellular cytokine production; Smac mimetic-based boosting was less likely to induce immune activation. We found that HIV-1 reservoirs in PWH contain unspliced and polyadenylated (polyA) virus mRNA, the ratios of which are greater in resting than total CD4^+^ T cells and can correct to 1:1 with PMAi exposure. Latency reversal results in greater fold-change increases to HIV-1 poly(A) mRNA than unspliced message. Multiply spliced HIV-1 transcripts and virion production did not consistently increase with LRA boosting, suggesting the presence of a persistent post-transcriptional block. LRA boosting can be leveraged to probe the mechanisms of an effective cellular HIV-1 latency reversal program.

## Introduction

HIV-1 proviral DNA remains integrated in CD4^+^ T cells during suppressive antiretroviral therapy (ART) and is a barrier to cure. This virus reservoir, defined as cells that contain provirus capable of generating infectious virions, is initially seeded within days of infection before symptoms develop (1, 2), persists (3-5), decays slowly (6), and can be maintained indefinitely (7, 8). The best approach to pursue HIV eradication remains uncertain. One strategy aims to pharmacologically reactivate latent HIV transcription in the setting of suppressive ART, induce virus protein production in infected cells, and then clear the reservoir through immune-mediated mechanisms (9-11). HIV-1 latency reversal agents (LRA) have advanced into clinical trials as monotherapy but with modest results; transient increases in virus cell-associated RNA (caRNA) production may be observed with some LRA but does not translate into a decrease in reservoir size (12-22). HIV-1 latency may be characterized by transcriptional blocks that cannot be overcome by single LRA (23-26).

To address these limitations, novel classes and combinations of LRA have been studied (27). Epigenetic LRA such as histone deacetylase inhibitors, when combined with protein kinase C agonists or second mitochondria-derived activator of caspase (Smac) mimetics, have been reported under some conditions to synergistically increase HIV-1 transcription in CD4^+^ T cells isolated from virally suppressed participants with HIV (28-32). Generally, the sample sizes were small and evaluated LRA concentrations in the mid-nanomolar to micromolar range. The absolute amount of HIV-1 latency reversal reported with combination LRA, defined primarily as induced levels of HIV-1 caRNA, has been significantly less than that observed with the maximal HIV-1 reactivating combination of phorbol 12-myristate 13-acetate (PMA) and ionomycin (PMAi). A trade-off may exist between maximum synergy and maximum efficacy in combination LRA approaches (33).

While HIV-1 is generally accepted to be transcriptionally silent during suppressive ART, this latency may not be absolute. Studies demonstrate that short promoter proximal transcripts of less than 60 nucleotides may be generated in the absence of the HIV-1 trans-activator of transcription protein, Tat (34, 35). More recently, resting CD4^+^ T cells isolated from PWH were found to contain transcripts extending beyond the transcriptional pausing site, a minority of which may be full-length or spliced and polyadenylated (36). Total CD4^+^ T cells isolated from PWH may contain approximately sevenfold more proximal HIV-1 transcripts, of at least 185 nucleotides in length, than polyadenylated virus mRNA (23). HIV-1 cell-associated RNA is readily detectable in the blood and tissue of PWH on virologically suppressive ART (37-40).

People with HIV may be exposed to opioids, whether prescribed for chronic pain (41), prescribed for opioid use disorder (42), or injected as heroin (43). The pharmacology of opioids associated with substance use disorders may differ from opioid medications used to treat chronic pain. The µ opioid agonists, e.g. heroin, fentanyl, morphine, and methadone, can block IL-2 mRNA and protein production in activated T cells through inhibition of AP-1, NFAT, and NF-κB activation, transcription factors known to be positive regulators of HIV transcription initiation (44-46). The effects of buprenorphine, a partial µ opioid agonist and κ and δ opioid antagonist commonly used in the treatment of opioid use disorder, on HIV-1 latency and its reversal have not been explored. A recent finding suggested that people with HIV who use opioids may have diminished responses to latency reversal mediated by T cell receptor (TCR) agonism (47).

To investigate the mechanisms that control HIV-1 latency reversal during opioid use, we enrolled a cohort of thirty-six treated, virologically suppressed participants with HIV. We originally hypothesized that opioid use would limit HIV-1 latency reversal and vary as a function of opioid use type. We tested LRA combinations at concentrations lower than those typically assessed and identified the phenomenon of LRA boosting: synergistic HIV-1 transcriptional reactivation with small molecules that do not reverse latency as single agents. We corroborated these findings in an additional eleven participants from a separate cohort and identified key associations between potent LRA boosting, increases in histone acetylation, modulation of T cell phenotype and function, and downstream blocks to virion production. While benchmarking LRA boosting, we observed that HIV-1 transcription during treated infection may be greater than previously appreciated and varies by CD4^+^ T cell phenotype. Importantly, we find that opioid use did not limit the magnitude or mechanisms of LRA boosting.

## Results

### Opioid use does not affect HIV-1 reservoir size

We enrolled the Opioids, HIV, and Translation (OPHION) cohort from a single academic center of thirty-six virally suppressed participants with HIV who used, or did not use, opioids in a 2:1 ratio. Participants had a median age of 59 years, included one-third women, and were approximately 30% white (**Table 1**). The median duration of ART was 12 years with a median duration of undetectable plasma HIV-1 RNA levels of over six years. Participant characteristics were further defined by type of opioid use (**Suppl. Table 1**) and validated by urine toxicology and self-reported substance use questionnaires (**Suppl. Fig. 1**). To reflect the range of opioid use encountered in clinical practice, this cohort comprised participants who actively injected opioids (n=4), used methadone (n=4) or suboxone (n=12) as medication for opioid use disorder, or took opioids for chronic pain (n=4). Amphetamine and barbiturate use were not detected. All active injection opioid users had confirmatory urine toxicology screens positive for fentanyl (n=2 of 4, range 62 - >500 ng/mL) and/or norfentanyl (n=4 of 4 participants, range 46 - >500 ng/mL) and reported a median of 6 days of injection opioid use in the preceding 30 days (IQR 4-16 days) with a median of 7 days since the most recent use (IQR 4-14 days). All participants in the methadone, buprenorphine, and prescription opioid subgroups had urine toxicology positive for that substance.

**Table 1.** OPHION Participant Characteristics.

| **Characteristic** | **Cohort** |  | **Total\*** |
| --- | --- | --- | --- |
|  | **Non-Opioid** | **Opioid** |  |
| Participants, N | 12 | 24 | 36 |
| Age |  |  |  |
| Median (IQR<sup>†</sup>) | 56 (50-61) | 61 (55-65) | 59 (54-65) |
| Sex |  |  |  |
| Male N (%) | 7 (58%) | 17 (71%) | 24 (67%) |
| Race |  |  |  |
| Black N (%) | 10 (83%) | 6 (25%) | 16 (44%) |
| White N (%) | 2 (17%) | 9 (38%) | 11 (31%) |
| Hispanic/Latino** N (%) | 0 | 2 (8%) | 2 (6%) |
| American Indian N (%) | 0 | 1 (4%) | 1 (3%) |
| Declined/NA | 0 | 6 (25%) | 6 (17%) |
| Ethnicity |  |  |  |
| Non-Hispanic N (%) | 11 (92%) | 13 (54%) | 24 (67%) |
| Hispanic N (%) | 1 (8%) | 11 (46%) | 12 (33%) |
| Duration of ART (months) |  |  |  |
| Median (IQR) | 162 (106-204) | 136 (91-173) | 144 (97-179) |
| Duration of viral suppression (months) |  |  |  |
| Median (IQR) | 69.5 (38-91) | 79 (51-94) | 75 (44-93) |

\*Percentage totals may not add up to 100 due to rounding. <sup>†</sup>IQR, interquartile range. \*\* Participants may identify as Hispanic/Latino either in response to a question about their race, or a follow-up question on ethnicity. Participants who reported Hispanic ethnicity identified their race as either “White” or “Declined/Not Available.”

To assess markers of HIV-1 persistence, we quantified total HIV-1 DNA and caRNA in PBMC and evaluated intact proviral reservoir size (**Fig. 1**). Total HIV-1 DNA levels were statistically similar between opioid and non-opioid groups, 2.36 vs 2.38 log_10_ copies/10^6^ PBMC, respectively, as were HIV-1 caRNA levels, 2.45 vs 2.52 log_10_ copies/10^6^ PBMC (**Fig. 1a**). Levels of intact provirus DNA were also similar between groups, 2.00 vs 1.82 log_10_ copies/10^6^

**Figure 1.**
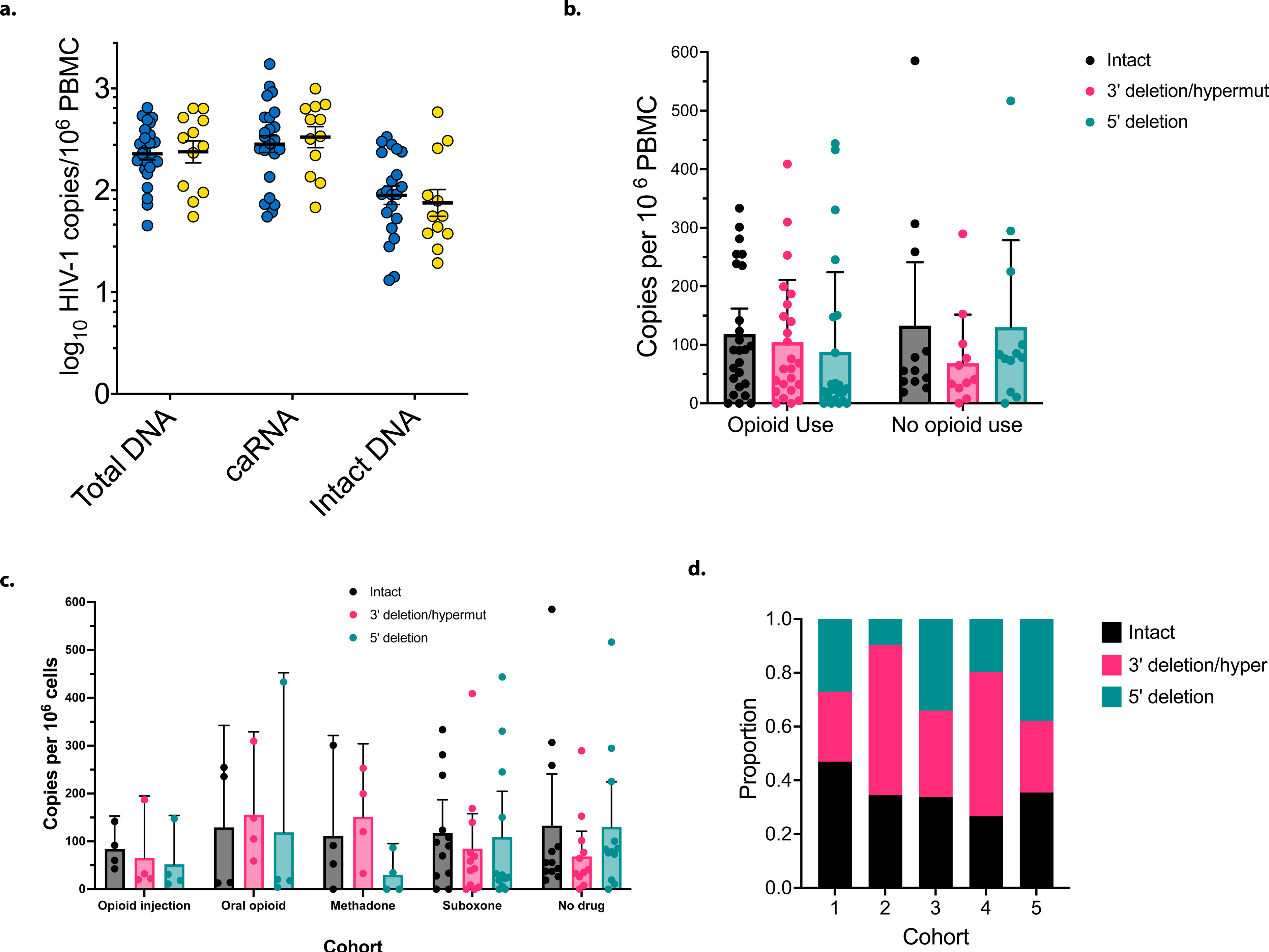
HIV-1 reservoir size and intactness during opioid use. Quantifications were performed on participants who used (n=24) or did not use (n=12) opioids. (a) HIV-1 persistence measures in participants with (blue circles) and without (yellow circles) opioid use are shown. Statistical significance was assessed with Wilcoxon signed-rank tests. Data are displayed on a logarithmic y-axis scale; DNA and RNA was extracted from 5 million PBMC. Intact DNA was quantified with the IPDA assay. Mean values ± standard errors of the mean (SEM) are shown. (b) 5’ and 3’ genomic deletions as a function of opioid use, mean values ± 95% CIs are displayed, (c) Opioid subgroup analysis. The reservoir size of intact, 5’ deleted, and 3’ deleted or hypermutated regions are shown. (d) The proportion of intact genomes as a function of DNA copies and opioid use subgroup; Kruskal-Wallis testing adjusting for multiple comparisons was not significant. Cohort 1, injection opioid use; cohort 2, prescribed oral opioids for pain; cohort 3, methadone; cohort 4, suboxone; cohort 5, no opioid use.

PBMC. To better define the intact proviral DNA reservoir, we characterized 5’ and 3’ HIV-1 deletions across cohorts and opioid use subgroups (**Fig. 1b-c**). Similar proportions of intact 5’ and 3’ genetic regions were observed, irrespective of opioid use. The ratios of proviruses containing an intact 5’ region and a defective (hypermutated) or deleted 3’ region in HIV-1 *env* were statistically similar. An exploratory analysis demonstrated a greater proportion of total genomes were intact in active injection opioid users, although this result was not significant when adjusted for multiple comparisons (**Fig. 1d**).

### LRA boosting markedly increases HIV-1 transcription

To determine the effects of opioid use on HIV-1 latency reversal *ex vivo*, we isolated PBMC from thirty-six participants enrolled in the OPHION cohort and tested a panel of ten LRA conditions (**Fig. 2**). The histone deacetylase inhibitors (HDACi) romidepsin (RMD) and panobinostat (PNB) were used at the lowest concentrations most commonly reported in the literature and the protein kinase C agonist bryostatin was tested at 1nM, one-tenth the most commonly reported concentration. In single LRA conditions, we observed a 2.7-fold activation of HIV-1 caRNA transcription, relative to an untreated DMSO-containing control, with anti-CD3/anti-CD28 (αCD3/αCD28) beads, a T cell receptor (TCR) agonist (**Fig. 2a**). This magnitude of TCR agonism-induced latency reversal is consistent with prior studies (23, 48-52). Statistically greater reactivation was observed with RMD (3.8-fold increase) when compared to PNB (2.6-fold increase) (**Suppl File 1**). Low-dose bryostatin minimally increased unspliced HIV-1 caRNA levels (1.2-fold increase), but the Smac mimetic AZD5582 (AZD), at a standard concentration (53), did not (0.9-fold increase, 95% CI [0.8, 1.1]).

**Figure 2.**
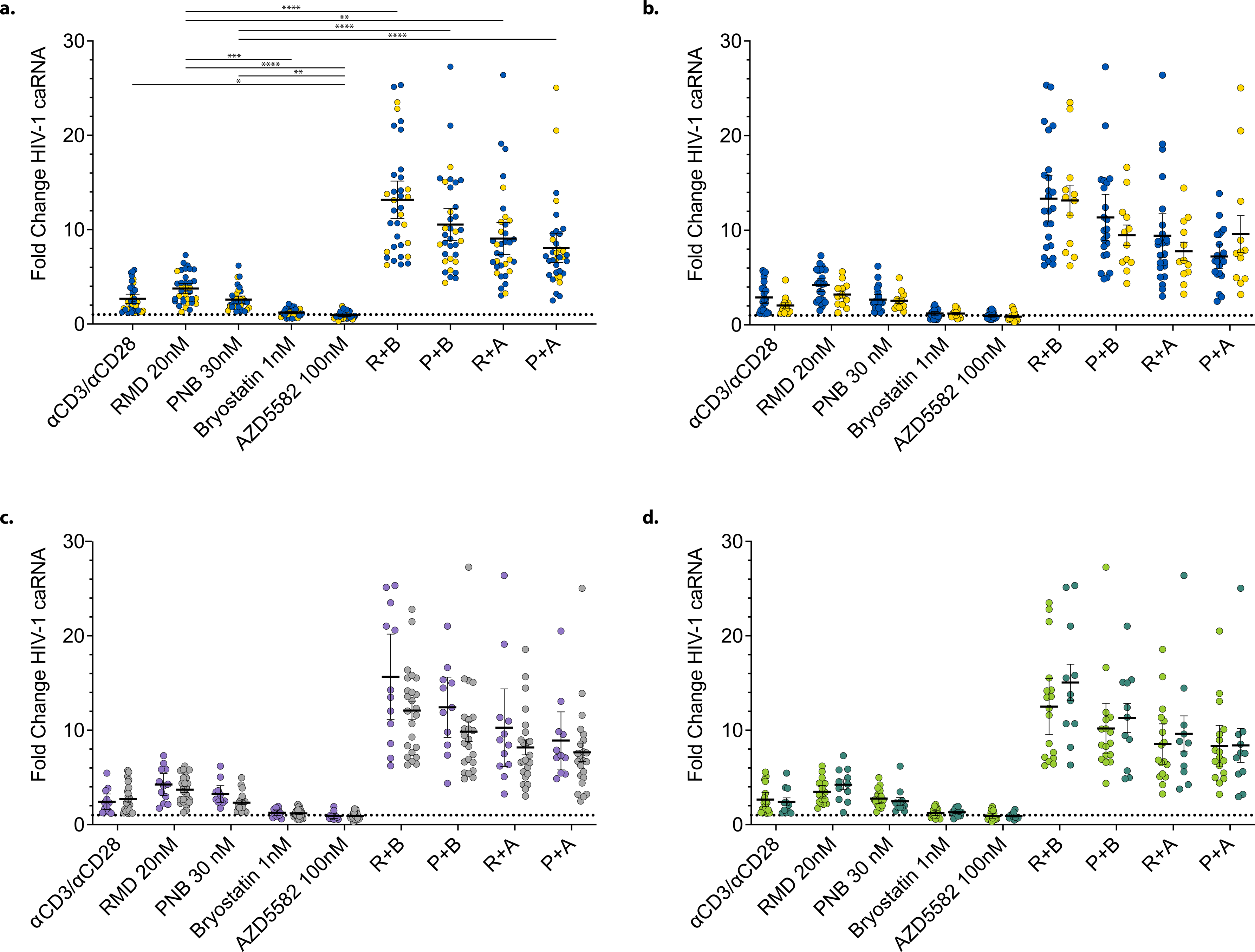
HIV-1 latency reversal in OPHION participants. HIV-1 caRNA levels were quantified after 24-hour incubations, in the presence of tenofovir 2 μM and raltegravir 2 μM, using 5 million PBMC and compared to a no-drug DMSO-containing condition. Mean values and the 95% CIs are shown. (a) Combined data showing LRA responses in thirty-six total participants with (blue circles) or without (yellow circles) opioid use. Data from (a) was dichotomized by (b) opioid use, (c) sex (light purple circles, male; light grey circles, female), and (d) race (light green circles, Black and American Indian; dark green circles, white). Means and SEM are shown. Dotted horizontal line denotes a fold-change of 1. *p<0.05, ** p<0.01, *** p<0.001, and **** p<0.0001 for LRA combinations, corrected for multiple comparisons. αCD3/αCD28, anti-CD3 anti-CD28 superparamagnetic beads; RMD, romidepsin; PNB, panobinostat; R, RMD 20nM; P, PNB 30nM; B, bryostatin 1nM; A, AZD5582 100nM.

We next assessed LRA in combination. Incubation of PBMC with HDACi and either AZD or low-dose bryostatin potentiated HIV-1 latency reversal. AZD in combination with PNB or RMD increased HIV-1 caRNA levels 8.1-fold and 9.1-fold, respectively. Low-dose bryostatin with PNB increased HIV-1 caRNA 10.6-fold, whereas the greatest fold-induction of HIV-1 caRNA was observed with the combination of low-dose bryostatin and RMD (13.2-fold). Statistically greater HIV-1 RNA induction was observed with the LRA boosting combinations relative to HDACi monotherapy and TCR agonism, when corrected for multiple comparisons. We compared LRA response as a function of opioid use (**Fig. 2b**), sex (**Fig. 2c**), race (**Fig. 2d**), and ethnicity (**Suppl. Fig. 2a**) and observed no statistically significant differences in the fold-changes of HIV-1 caRNA induction across these groups. In an exploratory analysis, the fold-change in HIV-1 caRNA varied by opioid use subgroup in response to T cell receptor agonism (**Suppl Fig 2b**). Statistically significantly greater reactivation with αCD3/αCD28 beads was observed in the suboxone sub-group when compared to active injection opioid users, the group with the smallest fold-change in TCR agonism-induced HIV-1 caRNA levels.

### LRA boosting modulates T cell activation and cytokine production

To understand whether LRA boosting activates CD4^+^ T cells, we assessed the upregulation of surface activation-induced markers (AIM) (54). We leveraged dual-marker AIM assays (**Suppl. Fig. 3, Fig. 3a**), originally developed to detect T cell antigen responsiveness, to quantify cellular activation that occurs with LRA exposure, outside the context of peptide-specific recognition. Using TCR agonism as a positive control, responses were detected in 49%, 32%, and 42% of CD4^+^ T cells in the OX40/PDL1, OX40/CD25, and CD69/CD40L AIM assays, respectively (**Fig. 3b**). Low-dose bryostatin significantly upregulated surface expression to more modest levels in all three AIM assays, relative to an untreated control condition. Bryostatin induced OX40/PDL1 surface expression in 5.2% of CD4^+^ T cells, levels of induction that did not significantly change when used in combination with RMD (5.4%) or PNB (3.5%). Bryostatin monotherapy induced expression in 6.9% and 2.6% of CD4s in the OX40/CD25 and CD69/CD40L AIM assays, respectively. In these two assays, the combination of RMD or PNB with low-dose bryostatin significantly reduced activation-induced markers, relative to bryostatin alone, and in some cases to control (untreated) levels; these findings did not differ by opioid use (**Suppl. Fig. 4a-c)**. The remainder of the LRA panel and their combinations did not upregulate surface expression in any AIM assay.

**Figure 3.**
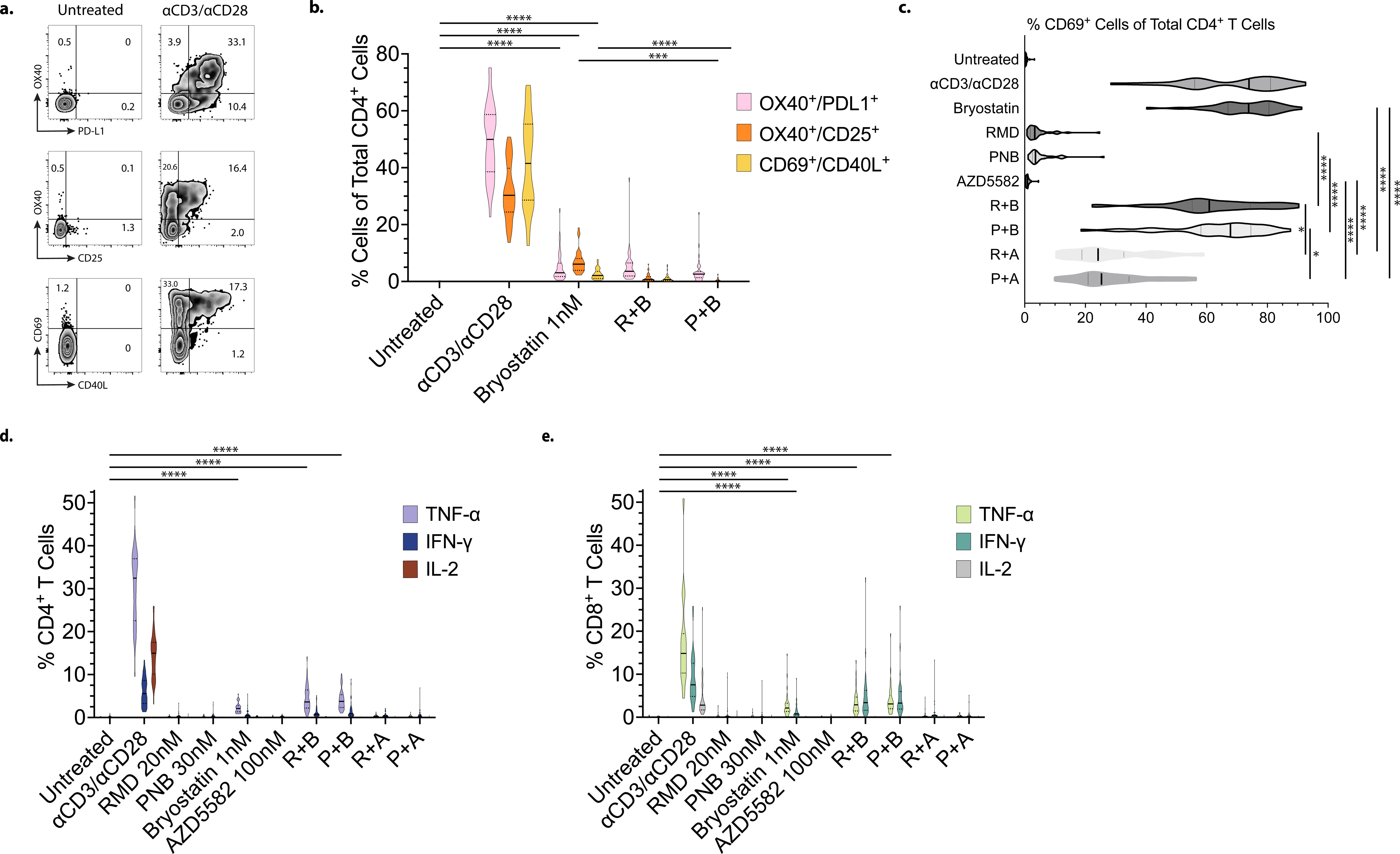
Immune activation and cytokine induction during *ex vivo* LRA boosting. (a) Example plots of OX40^+^PD-L1^+^, OX40^+^CD25^+^, and CD69^+^CD40L^+^ expression from gated live CD4^+^ T cells in PBMC samples incubated with DMSO control or an αCD3/αCD28 bead positive control for 18 hours, (b) Violin plots of surface activation-induced marker (AIM) response as a function of LRA. Results across three highly sensitive AIM assays for OPHION participants’ samples (n=36) are shown for comparison, (c) Quantification of LRA-induced surface CD69 expression in live total CD4^+^ T cells (n=36). Intracellular production of the cytokines TNF-α, IFN-γ, and IL-2 during LRA exposure in (d) live CD4^+^ and (e) CD8^+^ T cells (n=36). Violin plot median values are indicated by a solid black line; quartiles are shown with a dotted line. LRA conditions without visible bars represent values < 0.1% of cells. *p<0.05, *** p<0.001, and **** p<0.0001 for LRA combinations, corrected for multiple comparisons. RMD, romidepsin; PNB, panobinostat; R, RMD 20nM; P, PNB 30nM; B, bryostatin 1nM; A, AZD5582 100nM.

We observed a more pronounced upregulation in isolated CD69 expression, an early activation marker, in response to LRA. Approximately 0.5% of untreated CD4^+^ T cells expressed CD69 after 18 hours in culture (**Fig. 3c**). The proportion of cells expressing surface CD69 significantly increased to similar levels with TCR agonism (68.5%) and low-dose bryostatin (72.9%). Whereas AZD5582 (1.0%), RMD (4.7%), and PNB (5.4%) modestly increased CD69 levels, combinations of these small molecules led to synergistic effects. Significantly greater induction of CD69 expression was observed when RMD or PNB was combined with bryostatin or AZD5582; a bryostatin combination induced more CD69 than an HDACi plus AZD5582. CD69 levels were statistically similar when comparing TCR agonism, bryostatin alone, and bryostatin in combination with either HDACi (**Suppl. Fig. 4d**).

To further probe LRA effects on T cell activation, we stained CD4^+^ (CD4s) and CD8^+^ T cells (CD8s) for intracellular cytokine production (**Fig. 3d-e**). Whereas cytokine assessments in T lymphocytes most commonly include IL-2 and IFN-γ, we additional quantified intracellular TNF-α by flow cytometry. We observed a cytokine production hierarchy in CD4s, where the levels were TNF-α > IL-2 > IFN-γ, and in CD8s, where TNF-α > IFN-γ > IL-2. In response to TCR agonism, significantly more CD4s produce TNF-α (30.4%) than IL-2 (14.3%) or IFN-γ (6.0%), and significantly more IL-2 than IFN-γ (**Suppl. Fig. 4e**). We identified a different hierarchy in CD8^+^ T cells (CD8s), where the proportion of cells producing TNF-α remained greatest (17.5%) but significantly more IFN-γ (9.0%) was produced compared to IL-2 (4.3%). Whereas HDACi monotherapy did not increase the proportion of CD4s or CD8s producing cytokines compared to an untreated control, low-dose bryostatin alone or in combination with HDACi significantly increased production of TNF-α and, to a smaller magnitude, IFN-γ in CD4s. Significantly more CD4s produced TNF-α than IFN-γ in response to RMD + bryostatin (4.5% versus 1.3%) and PNB + bryostatin (4.2% versus 1.7%) (**Suppl. File 1**). In CD8s, low-dose bryostatin significantly increased production of TNF-α (2.8%) and IFN-γ (1.5%). The proportions of CD8s that produced TNF-α and IFN-γ when exposed to the combinations of RMD + bryostatin (3.4% versus 4.7%) and PNB + bryostatin (4.1% versus 4.6%) were similar and were statistically similar to bryostatin alone. IL-2 production did not increase above 0.1% of CD4s or CD8s for any LRA, or combination, we tested.

### LRA boosting increases histone acetylation

To investigate the mechanism(s) of LRA boosting, we next assessed histone acetylation. The proportions of acetylated histone H3^+^ live CD4s were similar between untreated cells (19.7%) and cells exposed to RMD (21.8%), PNB (21.7%), or AZD (19.8%) and significantly increased after single-LRA exposure to αCD3/αCD28 beads (35.5%) and low-dose bryostatin (29.2%) (**Fig. 4a**). Low-dose bryostatin in combination with either RMD (36.8%) or PNB (37.5%) significantly increased the proportions of CD4s contained acetylated histone H3. Interestingly, statistically significant increases in the proportions of acetylated histone H3^+^ live CD4s were also observed when combining LRA which individually did not affect these percentages, specifically RMD + AZD (30.0%) and PNB + AZD (29.8%). These results did not differ by opioid use (**Suppl. Fig 5a**).

**Figure 4.**
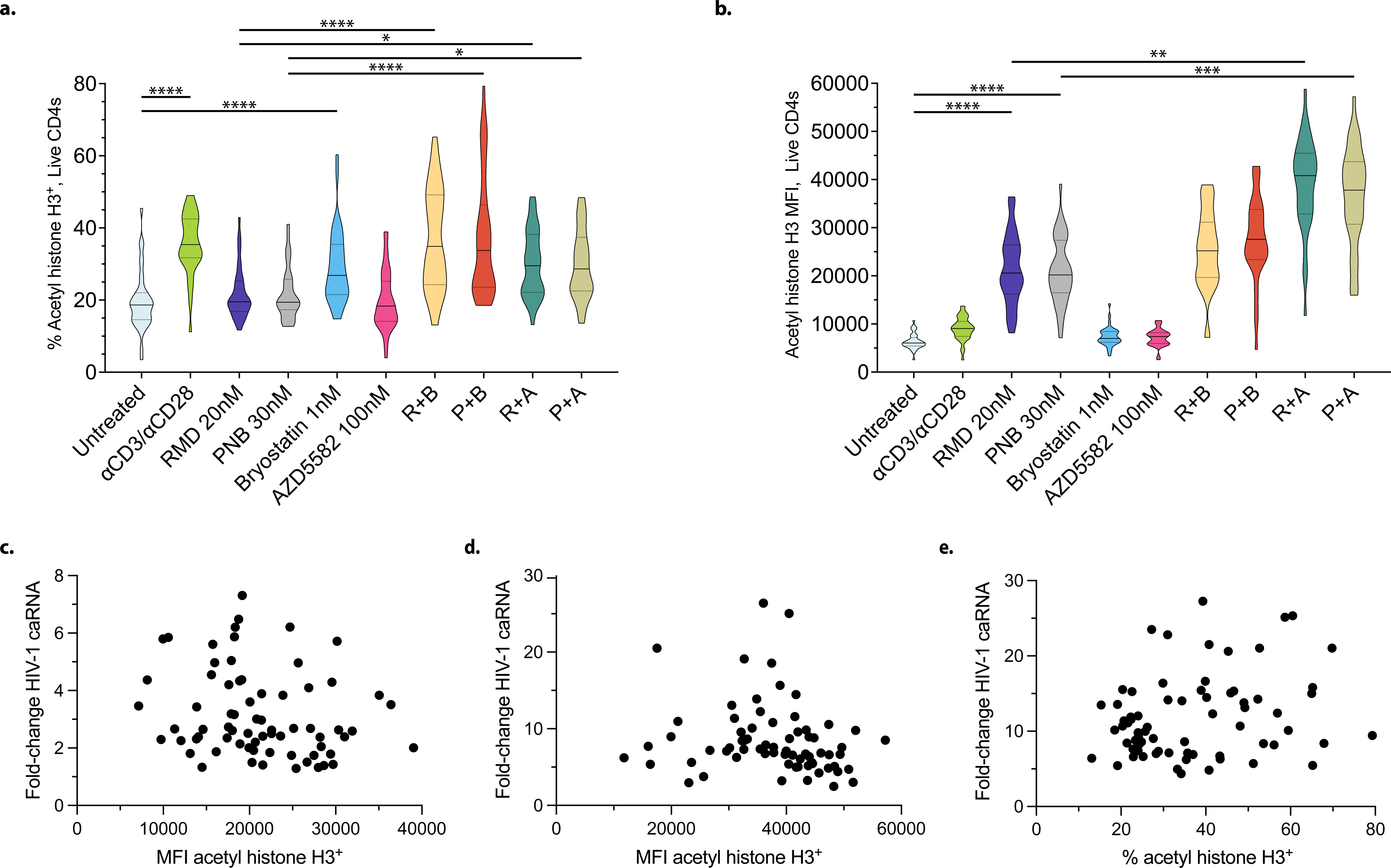
LRA boosting increases histone acetylation. (a) Violin plots showing the proportion of gated live total CD4^+^ T cells with acetylated histone H3, assessed by flow cytometry, (b) Violin plot of mean fluorescence intensity (MFI) of acetylated histone H3 per live CD4^+^ T cell. Scatter plots of acetylated histone H3 levels per CD4 as a function of HIV-1 transcription in response to RMD and PNB monotherapy (c), LRA boosting with HDACi and AZD5582 (d), and HDACi in combination with low-dose bryostatin (e) are shown. Acetylated histone levels and HIV-1 reactivation with the combinations of HDACi and bryostatin were moderately positively correlated (r=.25, p=.04). Violin plot median values are indicated by a solid black line; quartiles are shown with a dotted line.

Levels of acetylated histone H3, measured by median fluorescence intensity (MFI), were similar in untreated live CD4s and in cells cultured with αCD3/αCD28 beads, low-dose bryostatin, or AZD (**Fig. 4b**). Significant 3.4-fold and 3.3-fold increases in acetyl histone MFI were observed for RMD and PNB, respectively. AZD in combination with RMD or PNB further increased MFI by 6.1-fold and 5.7-fold, respectively. Absolute increases in MFI were observed for low-dose bryostatin in combination with RMD or PNB but did not retain significance after correction for multiple comparisons. These results did not differ by opioid use (**Suppl. Fig 5b**). We observed no association between acetylated histone H3 MFI and fold-change in HIV-1 RNA levels for HDACi monotherapy or for HDACi in combination with AZD (**Fig 4c-d**). A statistically significant weak association between the proportion of acetyl histone H3^+^ CD4s and fold-change in HIV-1 RNA levels was detected for HDACi in combination with bryostatin (**Fig 4e**).

### LRA boosting does not consistently induce virion production

To extend our analyses from the OPHION cohort, we next studied an additional eleven participants with HIV who did not use opioids, enrolled in the HEAL cohort, for whom leukapheresis samples were available (**Supplementary Table 2**). We compared LRA boosting responses in PBMC to the most potent known HIV-1 LRA, PMA in combination with ionomycin (PMAi). Significant fold-change increases in HIV-1 transcription were observed with combinations of low-dose bryostatin with RMD (8.3-fold) or PNB (8.0-fold), when compared to RMD alone (3.4-fold), PNB alone (3.1-fold), or PMAi (1.6-fold) (**Fig. 5a**). In separate experiments, we observed statistically similar HIV-1 transcription with an HDACi in combination with AZD, relative to PMAi (**Fig 5b**). No significant increases in HIV-1 transcription were observed in PBMC experiments with low-dose bryostatin (1.0-fold, 95% CI 0.9, 1.1) or AZD (1.2-fold, 95% CI 0.6, 1.9). Similar magnitudes of HIV-1 transcriptional reactivation were seen with HDACi monotherapy, low-dose bryostatin, and AZD monotherapy across the OPHION and HEAL cohorts we studied. We evaluated human reference gene transcription per million PBMC during LRA treatment and observed decreases with HDACi exposure, increases with PMAi, and variable changes with bryostatin (**Suppl. Fig. 6**) (37, 55).

**Figure 5.**
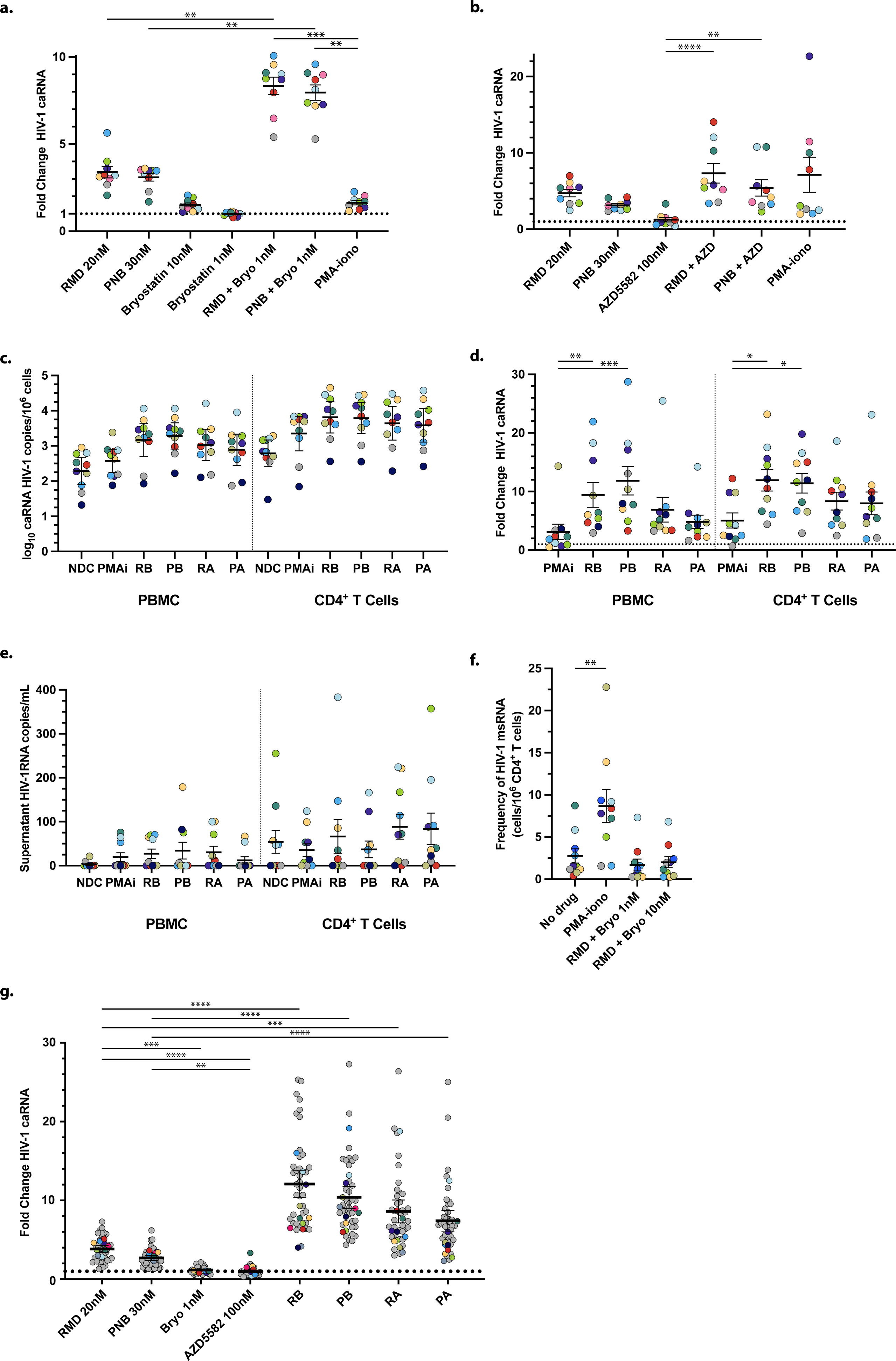
Comparative analysis of LRA boosting effects in PBMC and CD4^+^ T cells. Leukapheresis samples from HEAL cohort participants (n=11) were used to assess HIV-1 unspliced caRNA levels during (a) low-dose bryostatin-based and (b) AZD5582-based LRA boosting combinations in PBMC, when compared to PMA-ionomycin. Data are presented as means ± SEM. Circle color is used to denote a given HEAL participant across Figure panels. (c) Absolute HIV-1 unspliced caRNA levels in parallel PBMC and CD4^+^ T cell LRA exposures. PBMC and CD4 results are separated by a vertical dotted line. (d) Representation of the data in (c) as fold-change in HIV-1 caRNA levels. (e) Corresponding HIV-1 RNA levels from culture supernatants. (f) Multiply spliced HIV-1 RNA levels as determined in a modified TILDA assay, comparing RMD-bryostatin combinations at two bryostatin concentrations to PMA-ionomycin. (g) Combined OPHION and HEAL participant results (n=47) that summarize PBMC LRA boosting responses, for LRA conditions common to both datasets. Results from OPHION participant samples, carried over from Fig. 2a, are shown in grey circles. HEAL participant results are shown in color. *p<0.05, ** p<0.01, *** p<0.001, **** p<0.0001, corrected for multiple comparisons. NDC, no-drug control, contains 0.2% DMSO; PMAi, PMA with ionomycin; RMD, romidepsin; PNB, panobinostat; R, RMD 20nM; P, PNB 30nM; B, bryostatin 1nM; A, AZD5582 100nM.

To assess the magnitude of HIV-1 latency reversal as a function of cell type, we next performed parallel experiments in PBMC and total CD4^+^ T cells (**Fig. 5c**). While CD4^+^ T cells are the only established HIV-1 reservoir in PBMC, we performed this comparison across cell types to increase experimental rigor. Levels of HIV-1 caRNA were, on average, 4.3-fold higher (range 3.2 – 6.0) across experimental conditions performed in CD4^+^ T cells, when compared to PBMC, and varied by 1.6 log_10_ (PBMC) and 1.8 log_10_ (CD4) across participant samples in the absence of LRA exposure. To control for absolute caRNA differences, we determined fold-changes in LRA response and observed statistically significant increases in HIV-1 caRNA with low-dose bryostatin in combination with RMD or PNB, when compared to PMAi, in both PBMC and CD4s (**Fig. 5d**). To explore the variance in LRA response, we compiled per-participant biological replicate measurements (**Suppl. Fig. 7a**). Supernatant HIV-1 RNA levels increased after 24hr incubation of PBMC with PMAi (19 copies/mL, 95% CI [10, 32]) and the combinations of RMD plus bryostatin (27 copies/mL, 95% CI [11, 34]) or AZD (31 copies/mL, 95% CI [14, 43]), and PNB plus bryostatin (35 copies/mL, 95% CI [19, 60]) or AZD (12 copies/mL, 95% CI [8.1, 26]), when compared to untreated PBMC (3.0 copies/mL, 95% CI [2.2, 6.9]), but these results were not statistically significant after correction for multiple comparisons (**Fig. 5e**). This virion production response was dichotomized by participant. For each LRA condition, PBMC samples from at least 5 of 10 participants showed no increases in HIV-1 RNA levels over control; these were most commonly the same participants. Additionally, no significant increases in LRA-induced virion production in isolated CD4^+^ T cell cultures were observed. Greater virion production in untreated CD4s and greater absolute magnitudes of LRA-associated supernatant viremia in CD4s were noted, relative to PBMC, but these increases were seen in only a subset of participants. To define this post-transcriptional block more precisely, we quantified the frequency of CD4s with inducible multiply spliced HIV-1 RNA transcripts (**Fig. 5f**) (56). No significant increases in the frequency of cells that contain HIV-1 *tat*/*rev* transcripts were observed with RMD in combination with bryostatin at either 1nM or 10nM concentrations. In a post-hoc analysis, we combined datasets from OPHION and HEAL participants, including LRA conditions common to both sets of experiments, and observed statistically significant increases in HIV-1 unspliced RNA transcription with all LRA boosting combinations (**Fig. 5g, Suppl. Fig 7b**). The variance in LRA response, defined by the interquartile range, increased as the magnitude of fold change in HIV-1 caRNA increased.

### Effective latency reversal normalizes unspliced:polyadenylated HIV-1 mRNA ratios

We observed lower PMAi-associated fold-changes in HIV-1 caRNA levels with our unspliced HIV-1 RNA qPCR assay when compared to prior studies that quantified poly-adenylated (poly(A)) HIV-1 mRNA (28, 32). These assays differ methodologically and so we directly compared them and assessed the effects of primer location, CD4^+^ T cell phenotype, and CD4^+^ T cells density on HIV-1 caRNA quantifications (**Fig. 6a**). In response to a 24-hour PMAi exposure, levels of HIV-1 unspliced caRNA increased 4.3 – 8.6-fold. These levels did not significantly differ between resting (rCD4, CD25^-^CD69^-^HLA-DR^-^) and total CD4^+^ T cell (tCD4) stimulations, nor when CD4^+^ T cells were cultured at densities of 5x10^6^ or 1x10^6^ cells/mL. Poly(A) HIV-1 caRNA levels increased 32-34-fold with the same PMAi stimulation and were statistically significantly greater than unspliced HIV-1 caRNA fold-changes for each input CD4^+^ T cell phenotype or cell density. While PMAi-induced fold-changes in HIV-1 RNA were similar between rCD4 and tCD4, we observed differences in the amounts of unspliced and polyadenylated message present in these untreated cell populations. In the absence of LRA treatment, significantly greater ratios of unspliced to poly(A) HIV-1 caRNA were seen in rCD4 (10.6-13.3), relative to tCD4 (2.8-3.5) (**Fig 6b**). The differences in these ratios were driven by differences in HIV-1 poly(A) transcript levels. While untreated rCD4 and tCD4 at 5 million or 1 million cells/mL contained similar absolute levels of unspliced HIV-1 caRNA transcripts (p=0.98 for each), untreated tCD4 demonstrated statistically greater copy numbers of HIV-1 poly(A) mRNA per million CD4s (p<0.005 at 5m cells/mL, p<0.001 at 1mL cells/mL, Freidman test (ANOVA) with Dunn’s correction for multiple comparisons; **Suppl File 1**). PMAi stimulation normalized unspliced:poly(A) ratios to 1.7-2.4 in rCD4 and 0.8-1.0 in tCD4s (**Fig 6c, d**). Significantly lower ratios of unspliced:poly(A) HIV-1 RNA were observed in reactivated tCD4, when compared to rCD4 (**Fig. 6e**).

**Figure 6.**
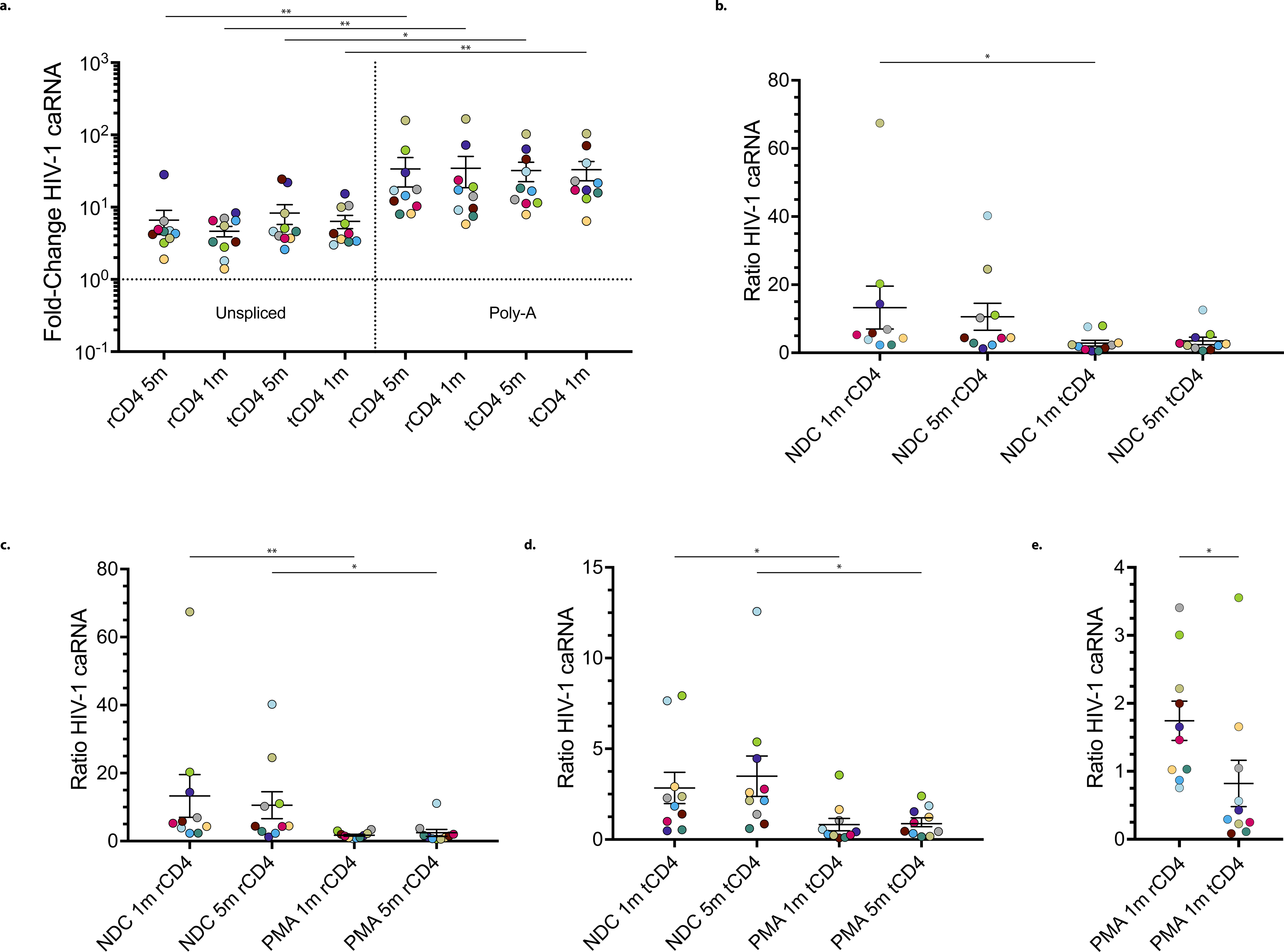
Latency reversal polyadenylates pre-existing unspliced HIV-1 mRNA (a) Resting and total CD4^+^ T cell HIV-1 transcriptional profiles during PMA-ionomycin exposure. HEAL participant samples (n=10) were used to assess the effects of CD4^+^ T cell phenotype and cell culture density on fold-changes in HIV-1 unspliced and poly(A) transcripts during latency reversal with PMA-ionomycin. (b) Ratios of HIV-1 unspliced and poly(A) transcripts in resting and total CD4^+^ T cells isolated from PWH. The effects of PMA-ionomycin and cell density on HIV-1 latency reversal in (c) resting CD4^+^ T cells, (d) total CD4^+^ T cells, and (e) the specific ratios of HIV-1 caRNA production in rCD4 compared to tCD4 when plated at a density of 1 million cells/mL, are shown. A given HEAL participant is indicated by circle color. Ratios of HIV-1 caRNA are the ratios of unspliced HIV-1 caRNA to poly(A) HIV-1 mRNA. *p<0.05, ** p<0.01, corrected for multiple comparisons in panels a-d. Panel e was assessed with a Wilcoxon matched-pairs signed rank test. rCD4, resting (CD25-CD69-HLA-DR-) CD4^+^ T cells; tCD4, total CD4^+^ T cells; NDC, no drug control condition; PMA, PMA 50ng/mL with ionomycin 1μM; Poly-A, polyadenylated HIV-1 caRNA; unspliced, HIV-1 unspliced caRNA; 1m, 1million cells/mL culture media; 5m, 5 million cells/mL culture media.

## Discussion

An approach to cure HIV infection should be equally effective for all people. Here we explored how opioid use may affect our ability to reverse HIV-1 latency. Opioid use is complex and can be accompanied by comorbidities, such as immune activation with injection drug use (57, 58), that may impact the HIV-1 reservoir and its persistence (37, 59-62). Here we found no effects of opioid use by indication, type, or pharmacology on markers of HIV-1 persistence. In the setting of persistently suppressed viremia, opioid use was less likely to modulate virus reservoir size, findings that agree with recent studies of injection heroin use (47, 63). Overall, participants were matched on the duration of ART and virus suppression, key variables that have been associated with HIV-1 reservoir size (64), and our urine toxicology results were consistent with the participants’ ascribed opioid use group.

We used samples from opioid users to identify the phenomenon of LRA boosting: improving the potency of HDACi beyond that of PMA-ionomycin by combining with a 2^nd^ drug that does not, by itself, re-activate HIV-1 transcription. This boosting occurred at lower LRA concentrations, here at least half the RMD, 1/10^th^ the bryostatin, and 1/100th the Smac mimetic concentrations used in prior combination studies (28-31). Synergistic latency reversal effects of low-dose bryostatin (1nM) and romidepsin was previously identified in samples from four participants with HIV (28). Here we leveraged samples from forty-seven participants to confirm those findings, extend the observation to HDACi in combination with low-dose bryostatin more generally, and to LRA boosting with a Smac mimetic. A recent study identified that injection opioid use was associated with a lack of ex vivo HIV-1 activation to TCR agonism (47). and our findings in the subgroup of active injection opioid users agree with this. However, this observation did not extend to other types of opioid use, e.g. suboxone use. Additional studies are needed to corroborate these findings in a larger sample size and investigate why HIV-1 latency reversal by TCR agonism may be particularly affected by injection opioids.

While AZD5582 has been reported to induce virus transcription, we observed no HIV-1 latency reversal activity when used as a single agent in PBMC isolated from the 47 participants we tested. AZD5582 monotherapy has been assessed for human ex vivo HIV-1 latency reversal activity in four prior studies, using the endpoints of a quantitative virus outgrowth assay, HIV-1 cell-associated RNA levels, and HIV-1 p24 production. In these studies, QVOA and p24 responses were not observed in the majority of participants, HIV-1 *gag* caRNA increases were approximately 2-fold, and the statistical significance of the results was less clear (53, 65, 66). Our findings agree more with a recent study that assessed AZD5582 monotherapy and did not identify HIV-1 LRA activity (32).

Our relatively large sample size, for this field, allowed us to determine that LRA boosting was less likely to be affected by sex, race, or ethnicity. We do note, however, that non-opioid participants in both our OPHION and HEAL cohorts were more likely to be black, whereas participants who used opioids were a more racially and ethnically mixed population. The use of two HDACi in our panel of ten LRA conditions increased the generalizability of the findings we observed with this drug class. Similar magnitudes of boosting were observed with RMD and PNB and suggests that class I histone deacetylase inhibition may be more relevant to this phenomenon (67, 68). HDACi-induced changes in acetylated histone (AcH) H3 and/or H4 are nearly exclusively reported in the literature as changes in MFI (13, 16-19, 69-72), not as a function of the percent AcH-positive cells (16). To increase transparency, we report histone acetylation changes both as the proportion of AcH-positive cells and by MFI; the 3-4-fold MFI increases we report with RMD and PNB are consistent with prior literature. LRA boosting correlated with increased histone H3 acetylation in CD4^+^ T cells but AZD-associated boosting did not recruit additional CD4s with acetylated histone H3. HDACi monotherapy also did not increase the proportions of acetylated histone H3-positive CD4s and the amount of acetylated histone H3 per cell did not meaningfully correlate with the induction of HIV-1 transcription. It remains unclear if histone acetylation is correlative or causative to virus reactivation. In the context of cellular genes, histone acetylation is not an immediate driver of the transcriptional changes observed with HDACi (73). Similar magnitudes of HIV-1 reactivation were observed with low-dose bryostatin and AZD5582-mediated HDACi boosting and could share common mechanisms. Bryostatin and AZD5582 can activate canonical and non-canonical NF-κB pathways, respectively, and the overlap in signaling with LRA boosting requires further definition. A recent study performed an AZD5582-stimulated CRISPR screen and identified HDAC2 as a potential synergistic drug target (32).

We identify that HIV-1 latency during treated virologically suppressed infection in PWH is best described a period of inefficient, but not absent, virus mRNA production in blood. Real-time PCR assays that quantify HIV-1 persistence markers are not necessarily standardized across research groups; the fold-change in HIV-1 unspliced caRNA levels that we observed with PMA-ionomycin stimulation were an order of magnitude lower than some prior studies which quantified polyadenylated HIV-1 mRNA (28, 32). Our findings suggest this is not a result of assay technical performance. Rather, qPCR assays of HIV-1 unspliced and poly(A) caRNA levels may capture distinct biology, as a function of CD4^+^ T cell phenotype. First, we identified tCD4 contain more polyadenylated HIV-1 message than rCD4 in PWH. Second, PMAi stimulation has more pronounced effects on the induction of HIV-1 poly(A) mRNA levels, relative to unspliced RNA, and explains the discrepancy in PMAi-induced HIV-1 caRNA fold-changes between our study and others. Third, PMAi stimulation increases the levels of both HIV-1 unspliced and poly(A) and, in doing so, normalizes their ratios; this normalization is greater in tCD4. Lastly, we identify that during treated virologically suppressed infection, HIV-1 transcription generates unspliced transcripts of at least 350 nucleotides in length. We note this is distal to where RNA polymerase II promoter-proximal pausing typically occurs (74). The efficiency of HIV-1 transcription is therefore reduced during ART, but not extinguished. We speculate that while the HIV-1 long terminal repeat (LTR) can recruit RNA polymerase II under these circumstances, the transcription elongation complex may be missing virus and/or host factors required to maximize virus mRNA production and processing.

LRA boosting, and their individual components, can have off-target effects. We used cell surface and intracellular assessments of immune activation to investigate whether LRA boosting can be uncoupled from immune activation. Whereas CD69 surface expression has been proposed as a biomarker of latency reversal potency in the context of PKC agonists, we found no correlation between CD69 upregulation and the magnitude of latency reversal (75, 76). We identified less induction of CD69 expression with RMD monotherapy than a prior study, despite similar staining and gating strategies (77). While low-dose bryostatin was sufficient to increase surface activation-induced markers and increase cytokine production in CD4^+^ and CD8^+^ T cells, albeit to relatively low levels, the increased HIV-1 reactivation seen with LRA boosting did not increase surface AIM and in some cases reduced it – specifically by adding an HDACi to bryostatin. However, intracellular cytokine production increased modestly with these same combinations and suggests that AIM assays and ICS staining provide complementary information in the assessment of investigational LRA. Our findings suggest that future investigations into LRA-indued cytokine production should include TNF-α quantifications, whose levels are more likely to increase than either of the more commonly assessed IL-2 and IFN-γ. In contrast, LRA boosting with AZD5582 resulted in relatively lower surface levels of CD69 and no induction of surface activation markers or cytokine production in T cells. Our results extend previous findings on the effects of combination LRA on CD8^+^ T lymphocytes to CD4^+^ T cells (78) and suggest that cytokine assessments of IL-2 production in CD4^+^ T cells provide the least yield in LRA evaluations. We conclude that LRA boosting approaches can affect the immune system differently and may work through different mechanisms. An LRA boosting approach that minimizes immune modulatory effects may be more appealing as an investigational intervention.

The variance in HIV-1 LRA responsiveness we document, across participants and between replicates, has implications for future studies. Small studies may be limited in their capacity to accurately quantify this variability, which increases with LRA potency and has been observed in other in vitro and ex vivo work (79-81). We speculate this variability may relate to heterogeneity in the HIV-1 reservoir, less so in the absolute amounts of provirus that may be found in a given PBMC aliquot and more so to the capacity of a given set of proviruses, in their unique intracellular environments, to support the reactivation of virus transcription. Assessments of LRA activity should prioritize larger sample sizes and more than one biological replicate per participant. We found that normalization of HIV-1 transcription to host gene transcription may introduce additional variability. Despite HIV-1 re-activation that was superior to PMA-ionomycin, multiply spliced HIV-1 transcription and virion production did not significantly increase with RMD in combination with low-dose bryostatin. Blocks to HIV-1 splicing are increasingly recognized, and here we extend those findings to novel LRA boosting combinations (23-25, 82-84). PMAi did not induce supernatant viremia, after a 24-hour incubation, in all participants; this heterogeneity has been previously reported (85). In CD4^+^ T cell samples from a subset of participants, however, we did observe detectable supernatant viremia but not increases in the frequency of HIV-1 msRNA with the LRA boosting combination of romidepsin and low-dose bryostatin. It is possible that if an RNA splicing block limits LRA boosting-associated virion production, it may not be absolute. Our study did not evaluate alternative possibilities that virion production without significant HIV-1 msRNA increases may be due to virion production kinetics, i.e. msRNA may be translated and degraded, may be due to the variance in LRA response we document across experimental replicates, or may be due to assay technical differences. Future work should focus on the post-transcriptional mechanisms of HIV-1 RNA processing, and inter-patient differences in LRA responsiveness.

To standardize language, the term HIV-1 “latency reversal” may require more precise wording. Latency reversal can refer to the reactivation of virus unspliced or poly(A) mRNA transcription but can also be interpreted to mean the reversal of latency that results in viral particle production. While some prior studies have identified HDACi-induced HIV-1 virion production *ex vivo*, this required HDACi exposures over 6-14 days, at times in combination with CD8 depletion or other LRA conditions that are more difficult to translate into investigational strategies (70, 77, 79, 86).

Our study has limitations. Prior and current injection opioid use limits the peripheral blood draw volumes we could obtain; this required us to prioritize the experiments we performed with a given sample. The statistical significance of LRA boosting with AZD5582-containing regimens was not replicated in our HEAL cohort samples and it is unclear if this relates to the variability in LRA response as sample size decreases or to unrecognized differences between our cohort’s participants. We observed variance across clinical cohorts in the magnitude of LRA boosting responses to AZD5582 in combination with HDACi. While the sample sizes for these experiments differ (n=36 vs n=9) and, in aggregate, the HIV-1 caRNA fold-change effects are clear, we cannot rule out that smaller sample sizes and/or unrecognized differences between our cohort’s participants may impact the significance and the magnitude of latency reversal with some LRA boosting combinations. Additional mechanisms may explain how LRA boosting increases virus transcription; bryostatin may exert its HIV-1 latency reversal effects independently of PKC agonism (87).

The use of HIV-1 LRA at sub-maximal doses and in combination with small molecules that do not increase virus transcription, *ex vivo*, expands our understanding of how virus latency may be reversed. Follow-on studies should delineate the precise mechanisms required for efficient virus RNA processing and virion production during HIV-1 latency reversal.

## Materials and Methods

### Study Design and Clinical Cohorts

The Opioids, HIV, and Translation (OPHION) study is a prospective cohort study of participants living with HIV on suppressive antiretroviral therapy recruited from Boston Medical Center which serves one of the largest populations of people living with HIV (PLWH) in Massachusetts, including a disproportionately large number of persons with opioid and substance use disorders. Eligible PLWH were required to currently be on antiretroviral therapy and virologically suppressed (defined as undetectable plasma HIV-1 RNA or <20 copies/ml) for a minimum of 12 months prior to enrollment. Individuals were recruited into 1 of 5 cohorts: (1) active injection opioid use, defined as self-report on injecting opioids at least once per week, (2) Methadone maintenance therapy for at least 3 months, (3) buprenorphine-naloxone therapy for at least 3 months, (4) Prescription oral opioid use for at least 6 months or (5) No opioid use within the past 1 year representing a control cohort. Eligibility into each of the five cohorts was confirmed with a urine toxicology performed at the BMC central laboratory, which includes evaluation for urine drug metabolites including opiates and non-opioid substances as well as an expanded opioid panel which included fentanyl or norfentanyl metabolites. Exclusionary criteria included ongoing immunosuppression and pregnancy; patients who reported injecting non-opioid substances were also not eligible. Once consented and enrolled, each participant underwent blood collection for same day PBMC extraction as well as urine collection for gabapentin and benzodiazepine metabolites. Participants also completed an extensive survey on their current and prior substance use (including alcohol and tobacco use) based the addiction severity index (ASI) (88) and Texas Christian University (TCU) drug screen (89). HIV and other clinical information including ART treatment history, duration of virologic suppression and comorbidity status including hepatitis C and B were abstracted from the medical record.

The HIV Eradication and Latency (HEAL) Cohort is a longitudinal biorepository study of participants with HIV. Inclusion criteria for HEAL participants in this study were 1) antiretroviral therapy-treated participants with HIV-1 who were virologically suppressed, defined as plasma HIV-1 RNA less than assay (most commonly less than 20 copies/mL, without blips) for >1 year and 2) had sufficient available amounts of cryopreserved PBMC for the proposed analyses. These cohort studies are performed in accordance with the principles of the Declaration of Helsinki. The institutional review boards at Boston Medical Center and Brigham and Women’s Hospital (Mass General Brigham Human Research Committee) approved the study protocols. Written informed consent was obtained from all participants. Experimenters were blinded to all clinical data and to OPHION subgroup assignments.

### Quantification of HIV-1 persistence markers

Genomic DNA and total RNA were isolated from approximately 5x10^6^ peripheral blood mononuclear cells (PBMC) or total CD4^+^ T cells (AllPrep DNA/RNA kit, Qiagen). Total HIV-1 DNA and unspliced cell-associated RNA (caRNA) levels were quantified in triplicate by real-time PCR as previously described, modified to include Taqman Universal Mastermix (DNA) or Taqman Fast Virus 1-Step Mastermix (RNA) (90). HIV-1 quantifications used forward primer 5’-TACTGACGCTCTCGCACC-3’, reverse primer 5’-TCTCGACGCAGGACTCG-3’, and probe 5’ FAM-CTCTCTCCTTCTAGCCTC-MGB 3’ (ThermoFisher Scientific). DNA cycling conditions in a total reaction volume of 25µL were 95°C for 15 min followed by 40 cycles of 95°C for 15 sec and 60°C for 1 min. RNA cycling conditions in a total reaction volume of 20µL were 55°C for 15 min, 95°C for 20 sec, followed by 40 cycles of 95°C for 3 sec and 60°C for 30 sec. Approximately 500ng of genomic DNA were assessed per well. The limits of quantification for total HIV-1 DNA and unspliced caRNA were 1 and 3 copies per reaction, respectively. Limits of detection were calculated per timepoint and participant sample, considering the average number of copies detected across triplicate measurements. Cell input numbers were quantified by human genome equivalents of CCR5 DNA using forward primer 5’-ATGATTCCTGGGAGAGACGC-3’, reverse primer 5’-AGCCAGGACGGTCACCTT-3’, and probe 5’ FAM-CTCTCTCCTTCTAGCCTC-MGB 3’ (ThermoFisher Scientific) as described (90).

### Intact proviral DNA assay

HIV-1 DNA from isolated PBMC DNA was measured using the intact proviral DNA assay (IPDA) as previously described with minor modifications (91). Two multiplexed droplet digital PCR assays (Bio-Rad QX200) were performed per sample targeting conserved HIV-1 sequences and, to normalize data, a cell reference gene. Master mixes were prepared using 2X ddPCR Supermix for probes. The HIV-1 specific reaction targeted the *Psi* packaging signal (Forward 5’-CAGGACTCGGCTTGCTGAAG-3’; Reverse 5’-GCACCCATCTCTCTCCTTCTAGC-3’; Probe 5’-TTTTGGCGTACTCACCAGT-3’, FAM, MGB) and *env* (Forward: 5’-AGTGGTGCAGAGAGAAAAAAGAGC-3’; Reverse: 5’-GTCTGGCCTGTACCGTCAGC-3’; Probe intact: 5’-CCTTGGGTTCTTGGGA-3’, VIC/HEX, MGB) sequences with 700 ng DNA per well in duplicate. An additional probe without a fluorophore targeting hypermutated *env* sequence (Probe hypermutated: 5’-CCTTAGGTTCTTAGGAGC-3’, Unlabeled, MGB) was used to exclude defective gene quantification. To estimate the number of cells per reaction and correct for sheared DNA, separate reactions targeting two regions of the *RPP30* gene, RPP30-1 (Forward: 5’-GATTTGGACCTGCGAGCG-3’; Reverse: 5’-GCGGCTGTCTCCACAAGT-3’; Probe: 5’-CTGACCTGAAGGCTCT-3’, VIC/HEX, MGB) and RPP30-2 (Forward: 5’-GACACAATGTTTGGTACATGGTTAA-3’; Reverse: 5’-CTTTGCTTTGTATGTTGGCAGAAA-3’; Probe: 5’-CCATCTCACCAATCATTCTCCTTCCTTC -3’, VIC/HEX), with similar nucleotide distance between targets compared to *Psi* and *env* were prepared with 70 ng DNA per well. Droplets were generated and subjected to PCR cycle: 95°C for 10 minutes, followed by 45 cycles of 94°C for 30 seconds, 59°C for 1 minute, and ending with 98°C for 10 minutes before holding at 4°C. Data from PCR products were collected and analyzed using QuantaSoft Data Analysis Software. Droplets containing a double positive signal (*Psi*+/*env*+) were considered intact, while single positive droplets represented defective provirus containing 5’ deletions (*Psi*-/*env*+) or 3’ deletions/hypermutations (*Psi*+/*env*-). Relative DNA shearing was calculated using the ratio of single positive RPP30 droplets compared to the double positive population and was used to estimate the number of intact proviruses. The number of diploid cells in each reaction was determined by dividing the copies of RPP30 in half and correcting for the dilution factor for the HIV-1 reaction. Intact, defective, and total HIV-1 DNA were normalized to the number of cells and reported as copies/million PBMC.

### Multiply spliced HIV-1 transcripts

To measure the frequency of total CD4^+^ T cells with inducible multiply spliced HIV-1 RNA, we performed the TILDA assay, as previously described, with two modifications (56). First, to reduce the 95% CIs around our frequency estimates, we used 36 x 10^6^ cells/mL as our first dilution, rather than the 18 x 10^6^ cells/mL used in Procopio, et al. Second, to harmonize our PMA-ionomycin concentrations with LRA experiments, we reduced the PMA concentration from its original 100 ng/mL to 50ng/mL in our TILDA assays. The duration of virologic suppression in our HEAL participants was similar to previously reported participants (56).

### Reference gene quantification

Extracted PBMC RNA was used to quantify transcription levels of three host genes, IPO8, TBP, and UBE2D2, typically immediately after the loading of HIV-1 caRNA qPCR plates was complete. IPO8 quantifications used forward primer 5’-CCTTTGTACAACAGAAGGCAC -3’, reverse primer 5’-TGCACGTCTCAGGTTTTTGC -3’, and probe 5’ FAM-TCCGCATAAATCCATTGATTCTGC -MGB 3’ (ThermoFisher Scientific); TBP quantifications used forward primer 5’-CAGTGAATCTTGGTTGTAAACTTGA -3’, reverse primer 5’-TCGTGGCTCTCTTATCCTCAT -3’, and probe 5’ FAM-CGCAGCAAACCGCTTGGGATTAT -MGB 3’ (ThermoFisher Scientific); and UBE2D2 quantifications used forward primer 5’-GTACTCTTGTCCATCTGTTCTCTG -3’, reverse primer 5’-CCATTCCCGAGCTATTCTGTT -3’, and probe 5’ VIC-CCGAGCAATCTCAGGCACTAAAGGA -MGB 3’ (ThermoFisher Scientific). To generate IPO8 standards, we cloned a fragment corresponding to nucleotides 3636-3707 of IPO8 mRNA (Sequence ID NM_006390.4) into a pCR4-TOPO vector (ThermoFisher Scientific, Cat. No. K458001). To generate TBP standards, a fragment corresponding to nucleotides 649-1037 of TBP mRNA (Sequence ID NM_003194) was cloned into a pCR4-TOPO vector. To generate UBE2D2 standards, a fragment corresponding to nucleotides 729-1164 of UBE2D2 mRNA (Sequence ID NM_181838.2) was cloned into a pCR4-TOPO vector. Reference gene RNA was synthesized using T3 Megascript Kit (ThermoFisher Cat. No. AM1333 and cleaned (RNease MinElute Cleanup Kit, Qiagen, Cat. No. 74204) before serially diluting across a range of 10^6^ – 10^3^ copies per well. After TBP and UBE2D2 standards were individually validated, TBP (T) and UBE2D2 (U) RNA were diluted separately and aliquoted together as one set of multiplex standards. T and U standards were validated to demonstrate overlap of individual and multiplexed standards curves. The assay was optimized to run all host gene standards and samples on one 96-well reaction plate in a total reaction volume of 20µL using cycling conditions of 55°C for 5 min, 95°C for 20 sec, followed by 40 cycles of 95°C for 3 sec and 56°C for 30 sec. Reference gene standards and samples were run in duplicate. IPO8, TBP, and UBE2D2 RNA copies per million PBMC were calculated by dividing copies per total sample by number of cells per sample, determined by CCR5 quantification, and normalized to 1x10^6^ cells. Fold changes were calculated by dividing the RNA copy number obtained per million PBMC for each LRA condition by the 0.2% DMSO control condition’s reference gene RNA copies/million PBMC.

### HIV-1 latency reversal

OPHION and HEAL participant peripheral blood mononuclear cells (PBMC) were isolated from large volume (90 – 180cc) peripheral blood or leukapheresis collections by density centrifugation and cryopreserved. Cryopreserved PBMC were thawed, pelleted, and transferred to RPMI media supplemented with 10% fetal bovine serum, 2 µM raltegravir, and 2 µM tenofovir (R10 + TDF/RAL) at a concentration of 1x10^6^ cells/mL. Cells were rested in a humidified CO_2_ incubator at 37°C for three hours (hrs). After 3hrs, cells were counted and assessed for viability. For CD4^+^ T cell experiments, PBMC were pelleted, resuspended in EasySep Buffer (StemCell Technologies, cat. no. 20144), and isolated with EasySep Human CD4^+^ T Cell Enrichment Kit (StemCell cat. no. 19052) per the manufacturer’s instructions. A total of 5x10^6^ PBMC, or total CD4^+^ T cells, were aliquoted into 5mL of fresh R10 + RAL/TDF in 6-well sterile cell culture plates. Cells were incubated in the presence or absence of the following LRA conditions: 0.2% DMSO, 1:1 αCD3/αCD28 beads (Life Technologies, cat. no. 11131D) to cells with 30 U/mL IL-2 (R&D Systems, cat. no. 202-IL-500), romidepsin 20 nM (Sigma, cat. no. SML1175), panobinostat 30nM (Sigma, cat. no. SML3060), bryostatin-1 1nM (Sigma, cat. no. B7431), AZD5582 100nM (ChemieTek, cat. no. CT-A5582), romidepsin 20 nM plus bryostatin-1 1nM, panobinostat 30nM plus bryostatin-1 1nM 100nM, romidepsin 20 nM plus AZD5582 100nM, panobinostat 30nM plus AZD5582 100nM, and phorbol 12-myristate 13-acetate (PMA) 50ng/mL (Sigma, cat. no. P1585) in combination with ionomycin 1μM (Sigma, cat. no. I9657). To provide a known positive control for subsequent immunology assessments, αCD3/αCD28 beads were used with OPHION samples. Cultures were incubated at 37°C for 24 hours. After 24 hours, cells were prepared for nucleic acid extractions.

### Comparison of qPCR assays after reactivation

CD4^+^ T cells were isolated from cryopreserved HEAL participant PBMCs. Total CD4s (tCD4) were isolated using a CD3+CD4+CD8-isolation kit (StemCell cat. no. 17952) and resting CD4s (rCD4) were isolated using a CD3+CD4+CD8-CD25-CD69-HLA-DR-isolation kit (StemCell cat. no. 17962). Cells were pelleted and transferred to RPMI media supplemented with 10% fetal bovine serum, 2 μM raltegravir, and 2 μM tenofovir (R10 + TDF/RAL) at a concentration of 5×10^6^ cells/mL. Cells were rested in a humidified CO_2_ incubator at 37°C for 3h and then counted and assessed for viability. A total of 5×10^6^ tCD4s or rCD4^+^ T cells were aliquoted into either 1mL or 5mL of fresh R10 + RAL/TDF in a 24-well sterile cell culture plate or 6-well sterile cell culture plate, respectively. Cells were incubated in either 0.2% DMSO or phorbol 12-myristate 13-acetate (PMA) 50ng/mL (Sigma, cat. no. P1585) in combination with ionomycin 1μM (Sigma, cat. no. I9657). Cultures were incubated at 37°C for 24 hours. After 24 hours, cells were prepared for nucleic acid extractions. RNA was eluted in a total volume of 65uL and DNA eluted in a volume of 100uL. For HIV-1 unspliced caRNA quantifications, 10uL of RNA were assessed in triplicate using primers and probe that bind the HIV-1 *gag* region (90). To calculate copies of HIV-1 caRNA per million cells, copy numbers were averaged and cell counts normalized to a CCR5 DNA reference. For polyadenylated HIV-1 mRNA quantifications, 10uL (approximately 1 million cell equivalents) of RNA were run in triplicate using primers and probe targeting the nef to poly-A region of HIV-1 mRNA as previously described (28). Our HIV-1 unspliced and poly(A) caRNA amplifications are both one-step qPCR assays that use an HIV-specific primer to synthesize cDNA. Standards were generated as previously described (92). For poly(A) HIV-1 mRNA quantifications, copies per million cell equivalents were calculated by initial cell count prior to reactivation. Both assays used Taqman Fast Virus 1-Step Mastermix in a total reaction volume of 20μL and cycling conditions of 55°C for 15 min, 95°C for 20 sec, followed by 40 cycles of 95°C for 3 sec and 60°C for 30 sec. Fold-change was calculated by dividing copies per million cells in the PMAi condition by copies per million cells in the 0.2% DMSO no drug control condition for each sample.

### Activation Induced Marker (AIM) assay

AIM assays were performed as previously described (54). Briefly, cryopreserved PBMC were thawed, washed, resuspended in R10, and rested for 3 hours at 37°C. Following the 3-hour rest interval, the appropriate number of cells were transferred to a 48-well plate and subsequently treated with CD40 blocking antibody (Miltenyi Biotec, cat. no. 130-094-133) for a final concentration of 0.5ug/mL for 15 minutes at 37°C. Cells were incubated in the presence or absence of our 10-LRA panel, as previously described. After an 18hr incubation, cells were harvested and stained for 50 minutes at 4°C with the surface staining monoclonal antibodies (mAb); (PD-L1-PE/Cy7 (Biolegend, cat. no. 329717), CD40L-PE (BD Biosciences [BD], cat. no. 561720), OX40-APC (BD cat. no. 563473), CD69-BV650 (Biolegend cat. no. 310933), CD3-BV605 (Biolegend cat. no. 317322), CD4-BV421 (BD cat. no. 562424), CD8-PerCp-Cy5.5 (BD cat. no. 560662), CD25-BUV395 (BD cat. no. 564034) and LIVE/DEAD Near-IR stain (ThermoFisher cat. no. L34975), washed, fixed and permeabilized (BD cytofix fixation buffer, cat. no. 554655), and then stained for 30 minutes at 4°C with intracellular mAb Acetyl-histone H3-Alexa Fluor 488 (Cell Signaling Technology, cat. no. 9683S) in 1x perm/wash buffer (BD, cat. no. 554723). The cells were washed and fixed (BD cytofix fixation buffer, cat. no. 554655) prior to flow cytometry analysis.

### Intracellular cytokine staining

Cryopreserved PBMC were thawed, washed, resuspended in RPMI + 10% FBS (R10), and rested for 3 hours at 37°C. Following the 3-hour rest interval, the appropriate number of cells were transferred to a 48-well plate and incubated for 2 hrs with our panel of 10 LRA. Following this 2-hour incubation period, GolgiPlug (BD, cat. no. 555029) and GolgiStop (BD, cat. no. 554724) were added to each condition at a concentration of 1:1000 and 6:10000 respectively, then incubated at 37°C for an additional 16 hrs. Cells were harvested, pelleted, resuspended, stained for 20 minutes at 4°C with surface staining mAb (CD3-BV605, CD4-BV421, CD8-PerCp-Cy5.5 and LIVE/DEAD Near-IR stain), fixed and permeabilized as described above, and then stained for 30 minutes at 4°C with the intracellular mAb TNF-α-PE/Dazzle-594 (Biolegend cat. no. 502946), IL-2-PE/Cy7 (BD cat. no. 560707) and IFN-γ-BV510 (Biolegend cat. no. 502544). PBMC were washed and fixed in (BD cytofix fixation buffer, cat. no. 554655) prior to flow cytometry analysis, as described above.

### Flow cytometry

Cells were acquired on a BD LSRFortessa using FACSDiva. Analysis was performed using FlowJo software versions 10.6.2 (Treestar, Versions 10 for Mac). Representative gating strategies can be found in the supplemental figures.

### Statistical analysis

Statistical analyses were performed using GraphPad Prism 9 for macOS. Means, medians, 95% confidence intervals, and standard errors of the mean were calculated. The pre-specified primary outcome of OPHION was a comparison of log_10_ PBMC HIV-1 caRNA levels between participants who use and do not use opioids. The pre-specified outcome for OPHION latency reversal experiments was the difference in HIV-1 caRNA levels between LRA treatment groups and across opioid use groups. To assess for changes in HIV-1 caRNA, we performed Kruskal-Wallis testing of all participants together for each of the LRAs with Dunn’s multiple comparison test, comparing the mean ranks of LRA conditions. An identical approach was taken with a series of exploratory analyses with OPHION participant samples that assessed histone acetylation, surface activation, and intracellular cytokine production, and with the exploratory statistical analyses performed on experiments with HEAL participant samples, unless otherwise specifically stated in the text.

## Data Availability

All data produced in the present study are available upon reasonable request to the authors

## Acknowledgements

This work is supported by the National Institutes of Health (NIH) R61 DA047038 (NL and AT) and R33 DA047038 (AT) and was facilitated by the Providence/Boston Center for AIDS Research (P30AI042853) and the Harvard University Center for AIDS Research (P30AI060354). The project described was further supported by Clinical Translational Science Award 1UL1TR002541-01 to Harvard University and Brigham and Women’s Hospital from the National Center for Research Resources. The content is solely the responsibility of the authors and does not necessarily represent the official views of the National Center for Research Resources or the National Institutes of Health.

## Author contributions

Conceptualization: AA, JT, KC, NL, AT

Methodology: AA, JT, KC, NL, AT

Investigation: TL, JB, AA, AO, KC, HC, HJ, KC

Visualization: TL, JB, AA, AO, KC, AT

Funding acquisition: NL, AT

Project administration: AA, SR, NL, AT

Supervision: AA, NL, AT

Writing – original draft: TL, JB, AA, AO, NL, AT

Writing – review & editing: TL, AA, NL, AT

## Declaration of interests

NL is now an employee of Moderna, Inc. AT has received remuneration from EBSCO Dynamed. Additional authors declare that they have no competing interests.

## Supplementary Figures Legends

**Supplementary Figure 1.** Substance use in the OPHION cohort. We assessed substance use by (a) clinical urine toxicology testing and (b) self-reported substance use questionnaires. Substance use results are separated by cohort groups by participant ID. Red squares indicate the presence of that substance in a urine sample; unavailable data were marked with “X” in a gray box. An expanded opiate panel was performed to assess for the presence of different opiate formulations. Color squares in panel b reflect any reported use by participants in the last 30 days. The numbers inside a given box indicate the number of days used this substance was reported to be used in the last 30 days. Gradient of color from blue to red indicate increasing report of use whereas white indicates no use. Concomitant use of alcohol, tobacco, marijuana, and gabapentin in the OPHION cohort was common. *all fentanyl measures were confirmed by follow-on quantification. Participants in the buprenorphine and methadone groups reported daily use of the respective medication in the 30 days prior, whereas participants taking opioids for chronic pain took opioids for 27-30 days in the 30 days preceding their enrollment in OPHION. 75% (N=3/4) of active injection users had urine toxicology screens positive for cocaine corresponding with self-report; all three reported smoking cocaine in the prior 30 days to specimen collection. One of twelve participants in the methadone use group and 2/12 in the non-opiate use group also has urine toxicology screens positive for cocaine. All active injection opioid users reported using multiple drugs on the same day in the last 30 days compared to 1/12 participants in non-opioid use group and 10/20 participants in methadone, buprenorphine and prescription opiate use groups combined. N=3/4 active injection opioid users reported smoking marijuana in the last 30 days. 6 and 8 participants about the 12 participants in opioid use groups had urine toxicology screens positive for benzodiazepines and gabapentin; all but 1 in both cases reported that this substance was prescribed. No participant had urine toxicology screens positive for amphetamines or barbiturates. All participants injecting opioids used tobacco in the preceding 30 days and, overall, 23 of 36 OPHION participants used tobacco within the month prior to their blood collection. 25% (N=6/24) of opioid use participants consumed alcohol in last 30 days compared to 58% (7/12) in the control group.

**Supplementary Figure 2.** HIV-1 LRA boosting response in the OPHION cohort by subgroup analysis. (a) HIV-1 fold-increase in unspliced RNA transcription plotted as a function of self-reported Hispanic (dark purple circles) or non-Hispanic (light blue circles) ethnicity. (b) Opioid-use subgroup analysis highlighting LRA response to αCD3/αCD28 beads from Main Fig 1a, as a function of opioid use subgroup. Means and SEM are shown. Dotted horizontal line denotes a fold-change of 1. ** p<0.01 for LRA combinations, corrected for multiple comparisons. αCD3/αCD28, anti-CD3 anti-CD28 superparamagnetic beads; RMD, romidepsin; PNB, panobinostat; R, RMD 20nM; P, PNB 30nM; B, bryostatin 1nM; A, AZD5582 100nM.

**Supplementary Figure 3.** Flow cytometry gating strategies. Representative intracellular cytokine staining plots for (a) control and (b) αCD3/αCD28 bead-exposed PBMC are shown. (c) Gating strategy for the three AIM assays. A representative sample exposed to TCR agonism is shown.

**Supplementary Figure 4.** LRA-induced immune activation as a function of opioid use. Results obtained with the (a) OX40/PD-L1, (b) OX40/CD25, and (c) CD69/CD40L AIM assays are reported for all ten conditions in the LRA panel, where samples from participants with (blue circles) and without (yellow circles) opioid use are shown. Data are represented as means ± 95% CIs. For the OX40/PL-L1 assay, the highest measured levels in OX40/PDL1 assay with bryostatin 1nM, R+B, and P+B are the same non-opioid-using participant. (d) Individual data points for CD69^+^ live total CD4+ T cells, as a function of opioid use. (e) Comparative hierarchies of intracellular cytokine production in CD4^+^ and CD8^+^ T cells exposed to TCR agonism (αCD3/αCD28 beads). Data displayed in these panels are identical to the data values shown in Main Fig. 3. *p<0.05, ** p<0.01, **** p<0.0001, corrected for multiple comparisons. TNF, tumor necrosis factor alpha; IFN, interferon gamma; IL-2, interleukin-2; CD4, CD4^+^ T cells; CD8, CD8^+^ T cells.

**Supplementary Figure 5.** Histone acetylation effects during LRA boosting, as a function of opioid use. Using the same data displayed in Main Fig. 4, here we show individual data points, labelled as samples from participants with (blue circles) and without (yellow circles) opioid use. (a) The proportion of live total CD4^+^ T cells with acetylated histone H3, assessed by flow cytometry, (b) Mean fluorescence intensity (MFI) of acetylated histone H3 per live total CD4^+^ T cells.

**Supplementary Figure 6.** The effects of LRA boosting on human reference gene transcription. Using leukapheresis samples from HEAL participants (n=10), we assessed and compared fold-changes in transcription of three host genes: (a) IPO8, (b) TBP, and (c) UBE2D2. Reference gene multiplexing was validated for (d) TBP and (e) UBE2D2 quantifications. The rationale to study these three reference genes is as follows. IPO8 transcription is used as an internal control by the AIDS Clinical Trials Group’s Virology Specialty Laboratories to measure RNA integrity in a dichotomous way. IPO8 Ct values >27 are used to suggest degradation of cellular samples in storage, and samples with IPO8 Ct >27 do not report HIV-1 RNA values. Recent work by Nancie Archin’s group at the UNC HIC Cure Center identified TBP and UBE2D2 as two of the more stable host genes during their LRA exposures, which included PMA, AZD5582, and HDACi, among a panel of eight LRA.

**Supplementary Figure 7.** The effects of LRA boosting on human reference gene transcription. Using leukapheresis samples from HEAL participants (n=9), we assessed and compared fold changes in transcription of three host genes: (a) IPO8, (b) TBP, and (c) UBE2D2. Reference gene multiplexing was validated for (d) TBP and (e) UBE2D2 quantifications. The rationale to study these three reference genes is as follows. IPO8 transcription is used as an internal control by the AIDS Clinical Trials Group’s Virology Specialty Laboratories to measure RNA integrity in a dichotomous way. IPO8 Ct values >27 are used to suggest degradation of cellular samples in storage, and samples with IPO8 Ct >27 do not report HIV-1 RNA values. Recent work by Nancie Archin at the UNC HIV Cure Center identified TBP and UBE2D2 as two of the more stable host genes during LRA exposures, which included PMA, AZD5582, and HDACi, among a panel of eight LRA.

**Supplementary Figure 8.** Variance in LRA response. (a) Magnitude of HIV-1 caRNA transcription fold-changes in biological replicate LRA experiments performed with HEAL leukapheresis samples, using the same data as shown in Main Fig 5a-c, (b) Box and whiskers plot displaying the data of Fig. 5g as the mean (solid black horizontal line), inter-quartile range (25^th^ – 75^th^ percentiles denoted as the borders of the vertical rectangle), and 5^th^-95^th^ percentiles (whiskers, error bars). Values below and above the whiskers are shown as individual data points. PMA-iono, PMA with ionomycin; RMD, romidepsin; PNB, panobinostat; R, RMD 20nM; P, PNB 30nM; B, bryostatin 1nM; A, AZD5582 100nM.

## Supplementary Tables

**Supplementary Table 1** – Breakdown of OPHION cohort into opioid use subgroups

**Supplementary Table 2** – HEAL Participant Characteristics

## Notes

### Competing Interest Statement

Athe Tsibris has received remuneration or lab funding from Gilead Sciences, Merck & Co, and EBSCO-Dynamed.
Nina Lin is an employee of Moderna, Inc.

### Funding Statement

This work is supported by the NIH R61 DA047038 and R33 DA047038 and was facilitated by the Providence/Boston CFAR (P30AI042853) and the Harvard University CFAR (P30AI060354. The project was further supported by Clinical Translational Science Award 1UL1TR002541.

### Author Declarations

This study was reviewed and approved by the Institutional review board at Boston Medical Center and the Massachusetts General Brigham Human Research Committee

### Summary of Updates

This version of the manuscript has been revised for style and emphasis, and to include new data on the relationship between HIV-1 unspliced and polyadenylated mRNA levels during treated suppressed infection in people with HIV, and during ex vivo latency reversal.

