## Supplementary figures and images for "HIV-1 latency reversal agent boosting is not limited by opioid use"

### Supplemental Figure 1

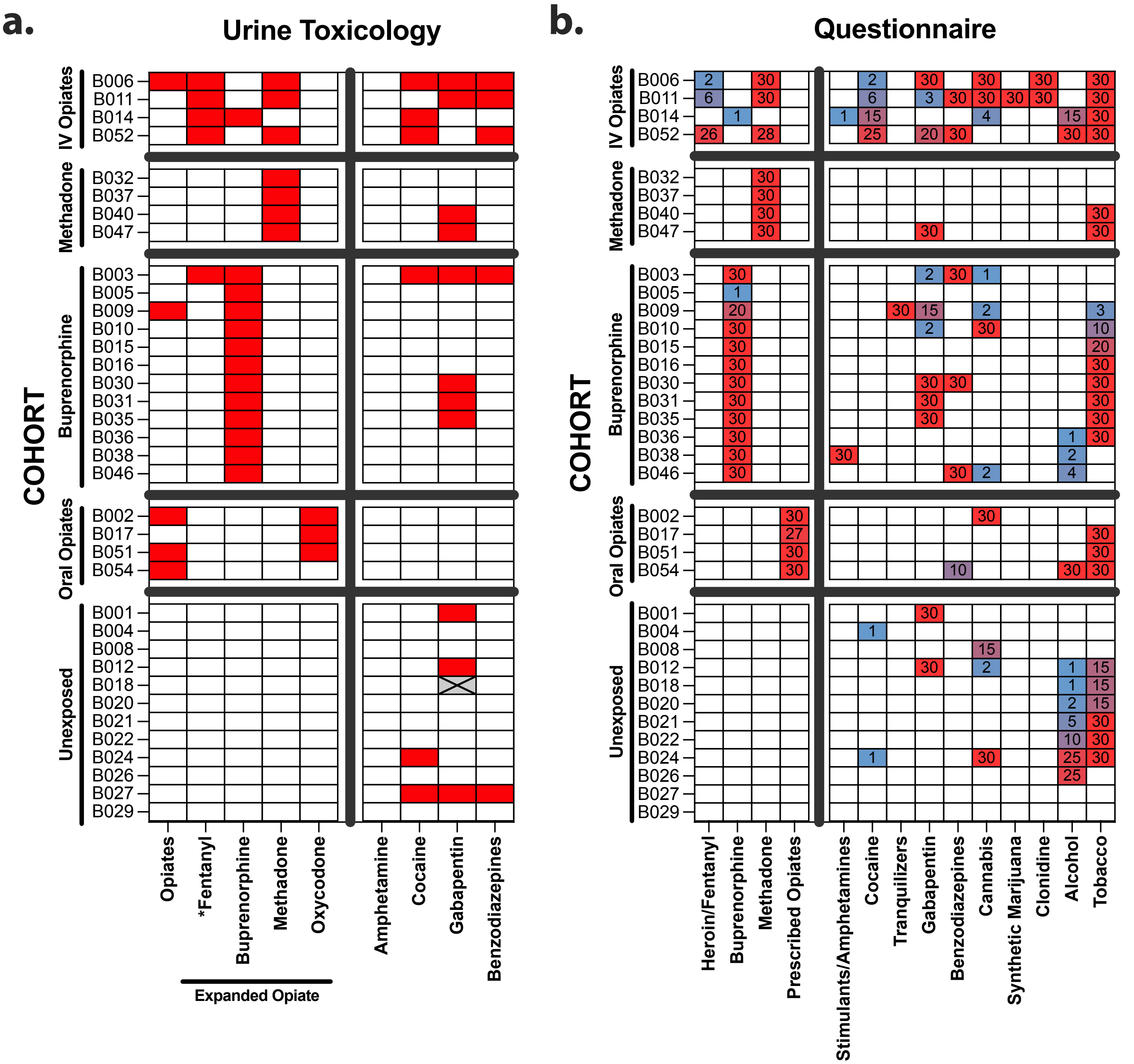

### Supplemental Figure 2

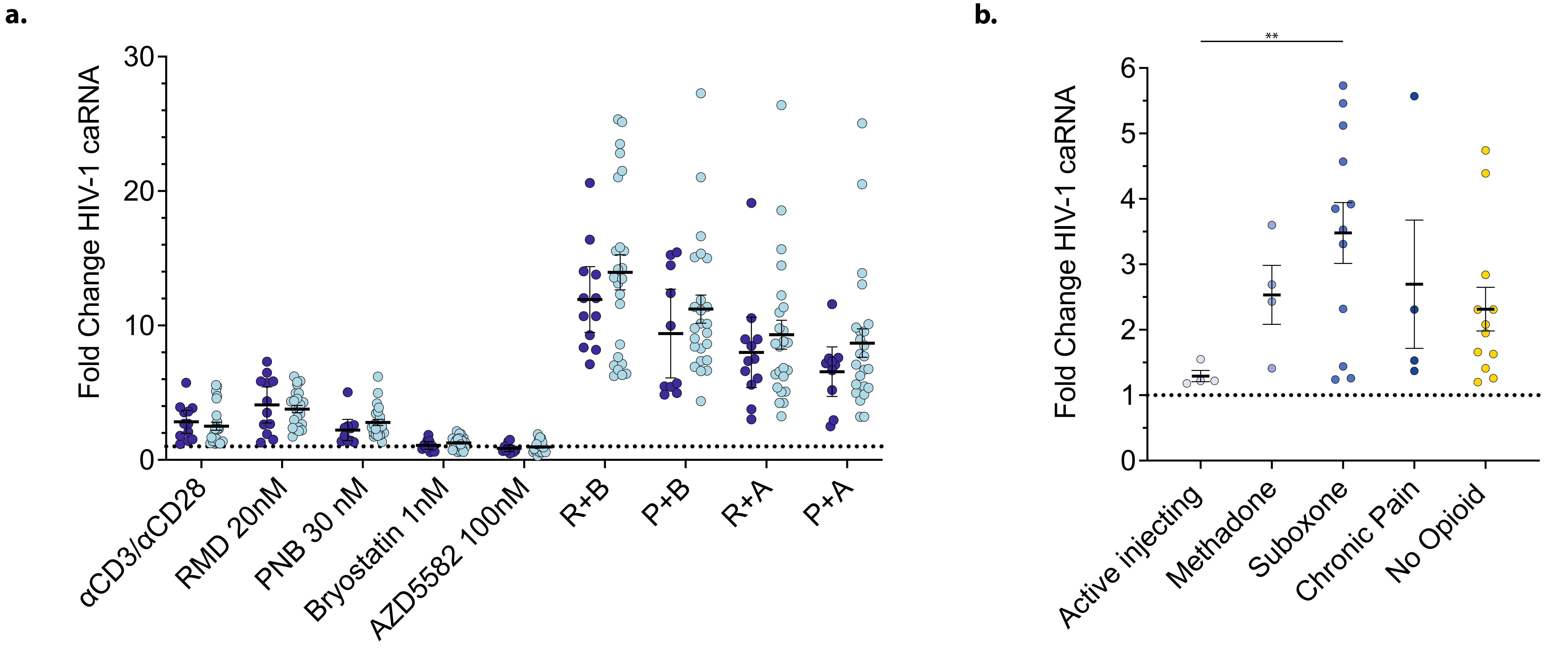


Supplementary Figure 2

### Supplemental Figure 3

#### Supplementary Figure 3

**a.**

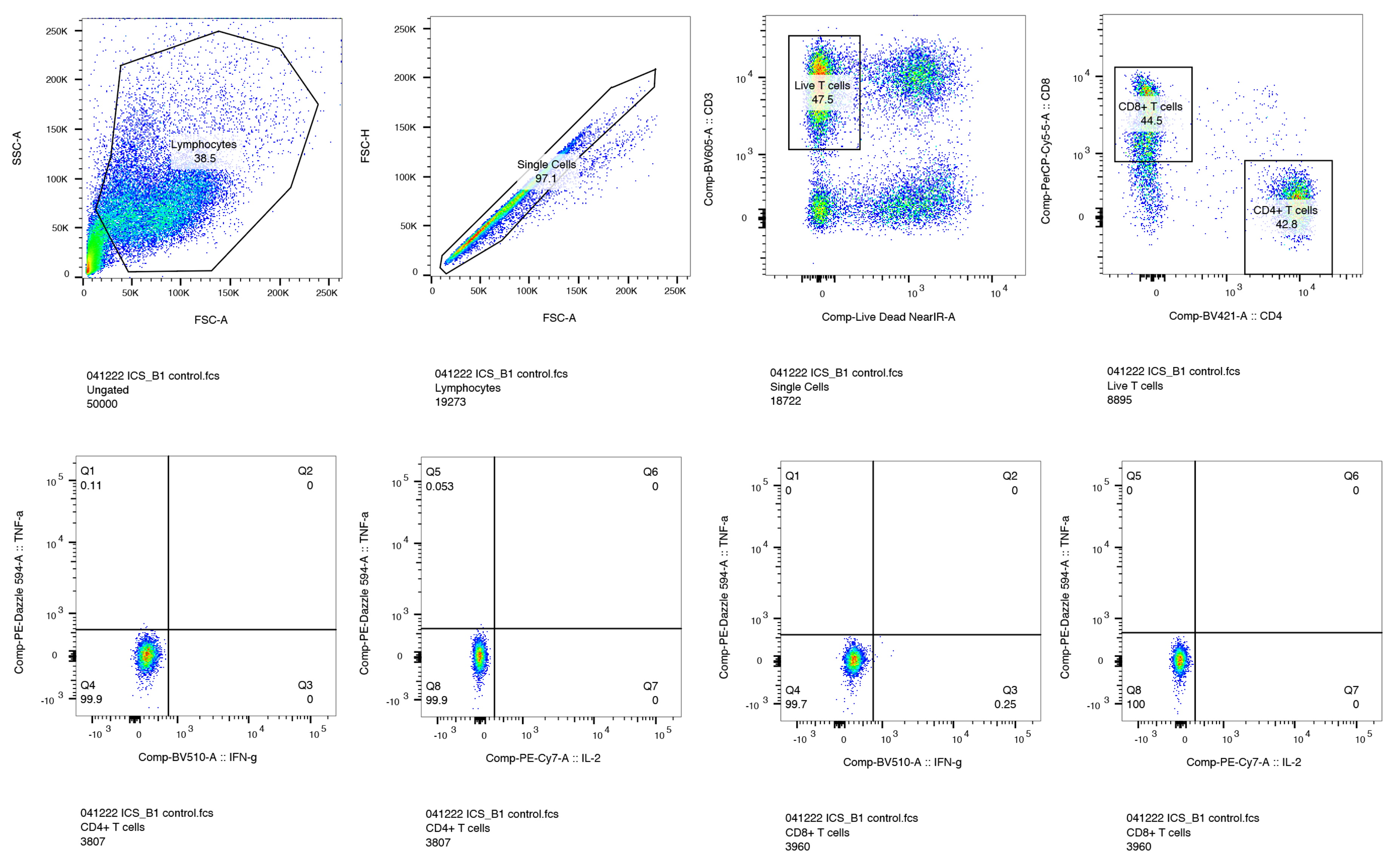

**b.**

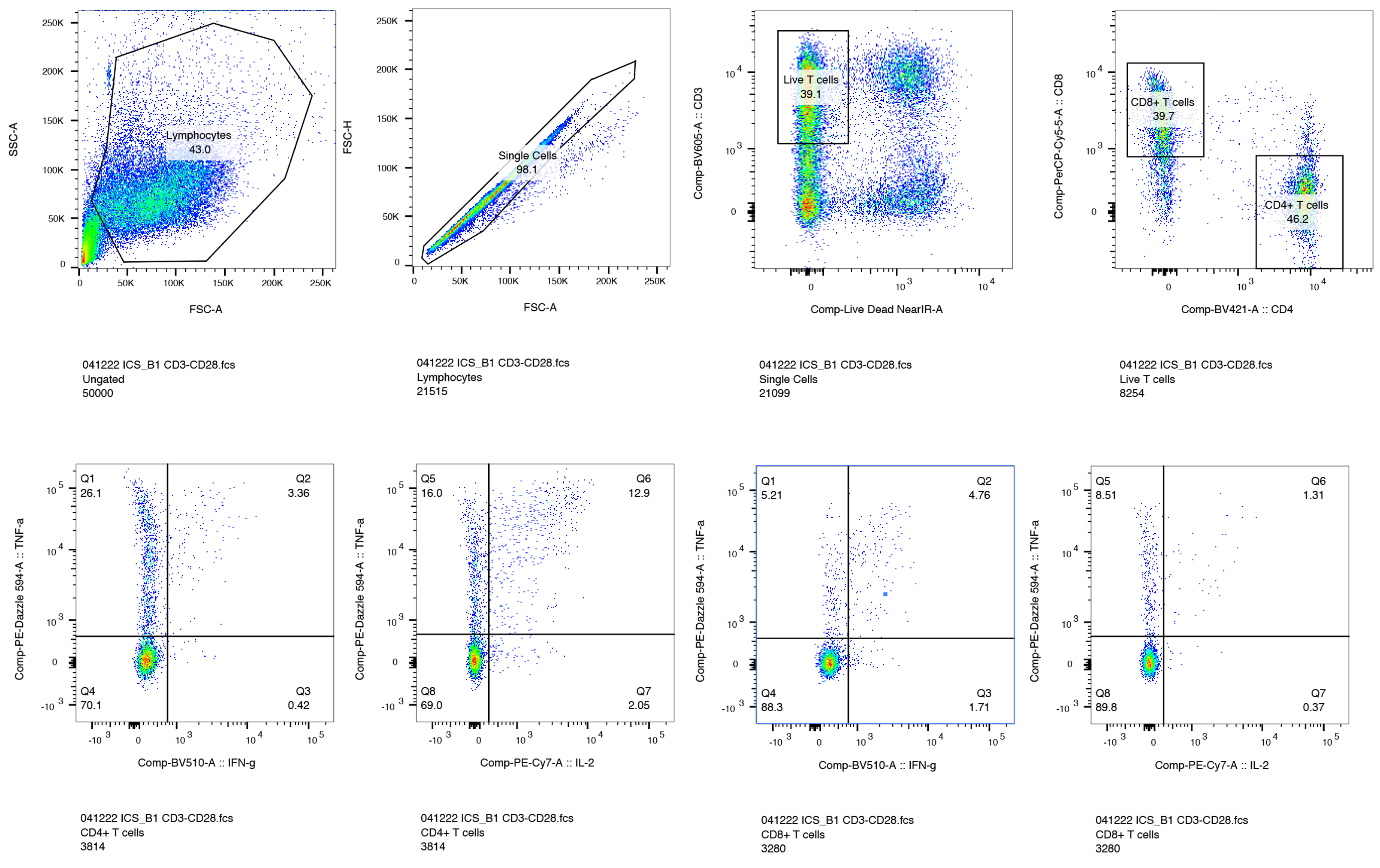

**c.**

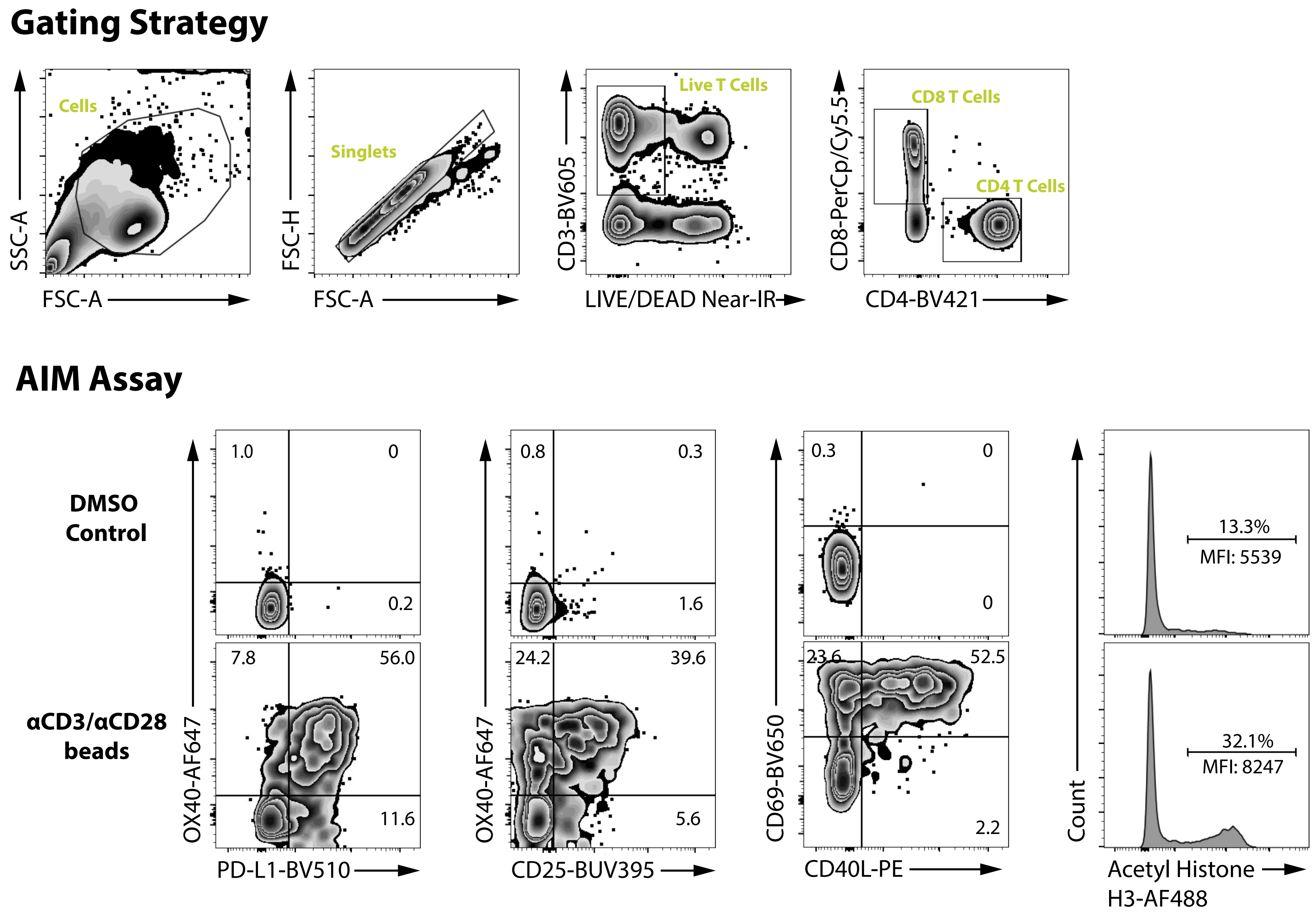

### Supplemental Figure 4

Supplementary Figure 4

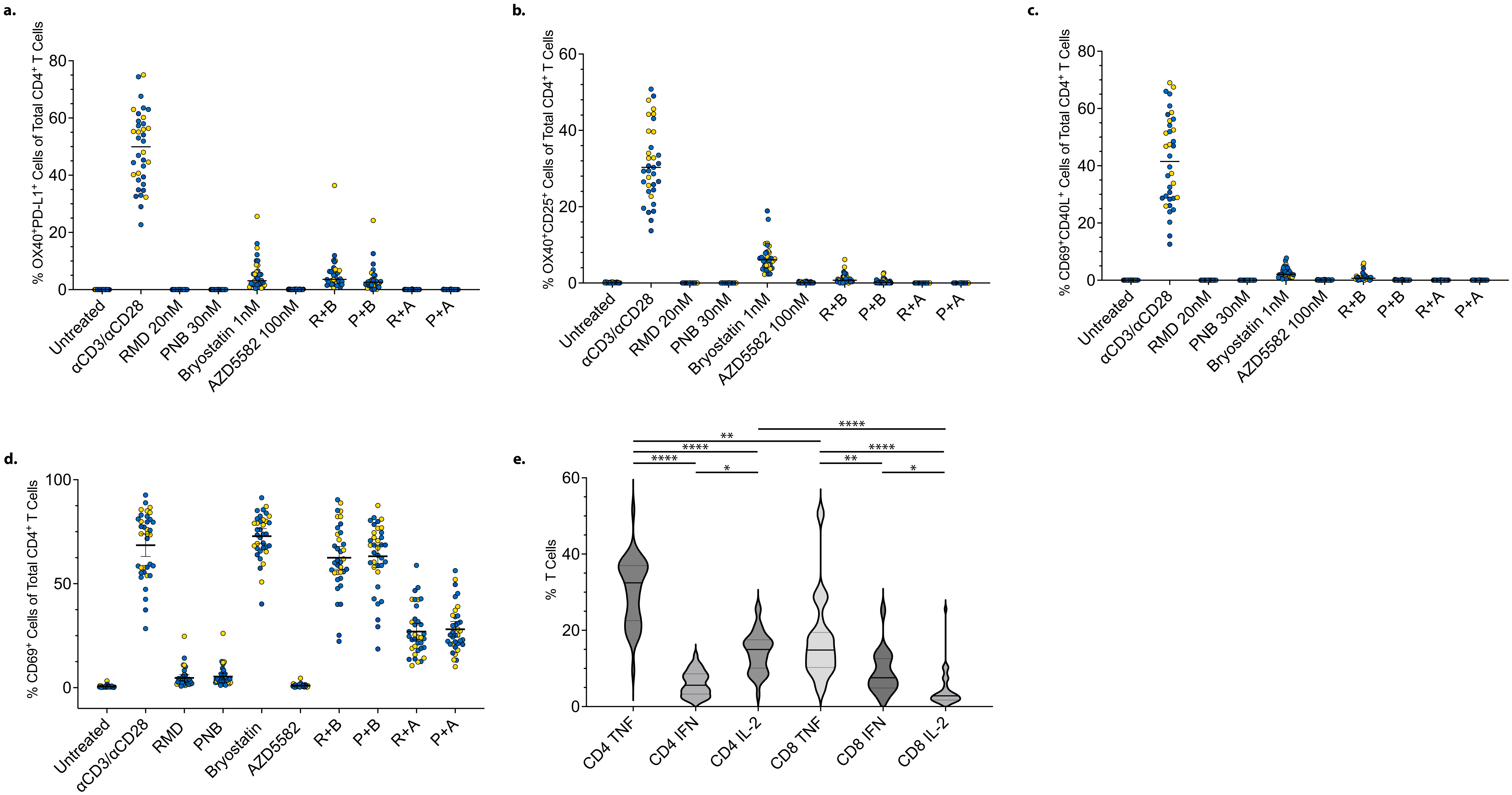

### Supplemental Figure 5

Supplementary Figure 5

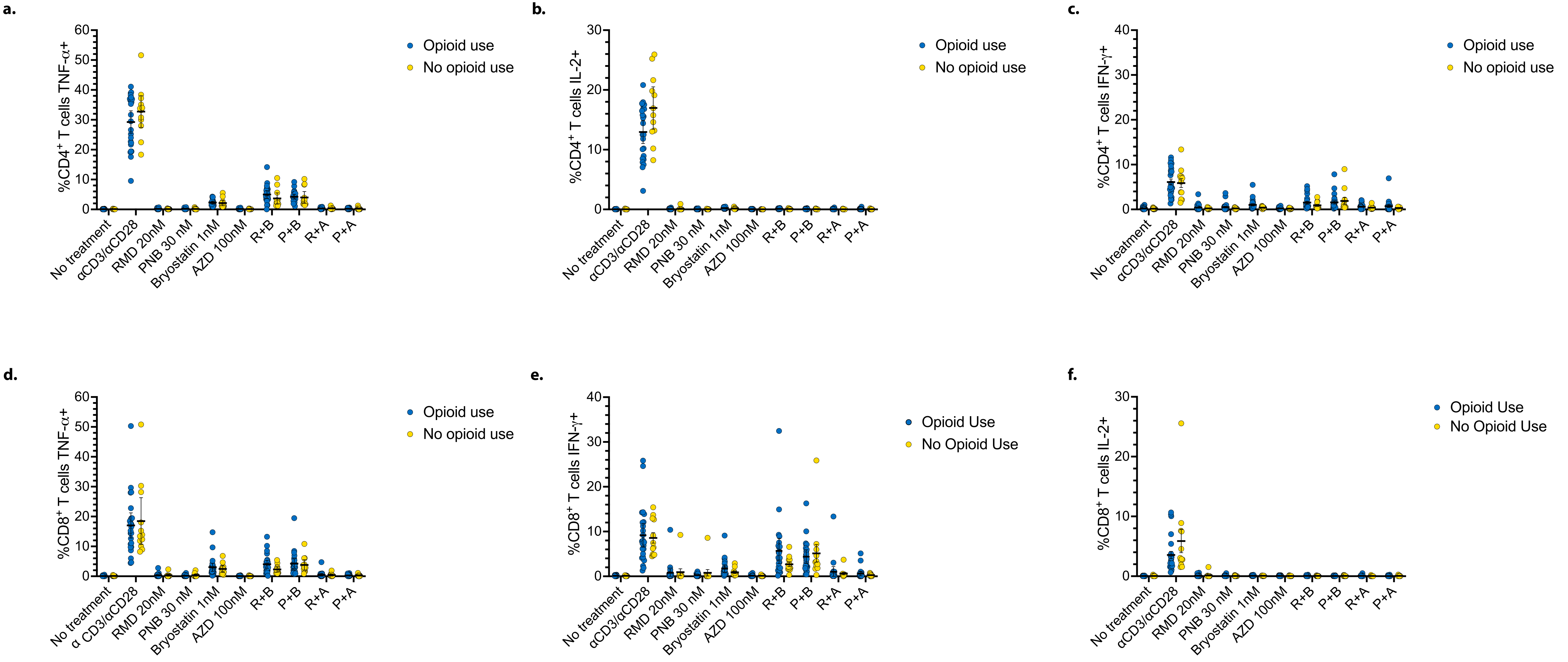

### Supplemental Figure 6

Supplementary Figure 6

**a.**

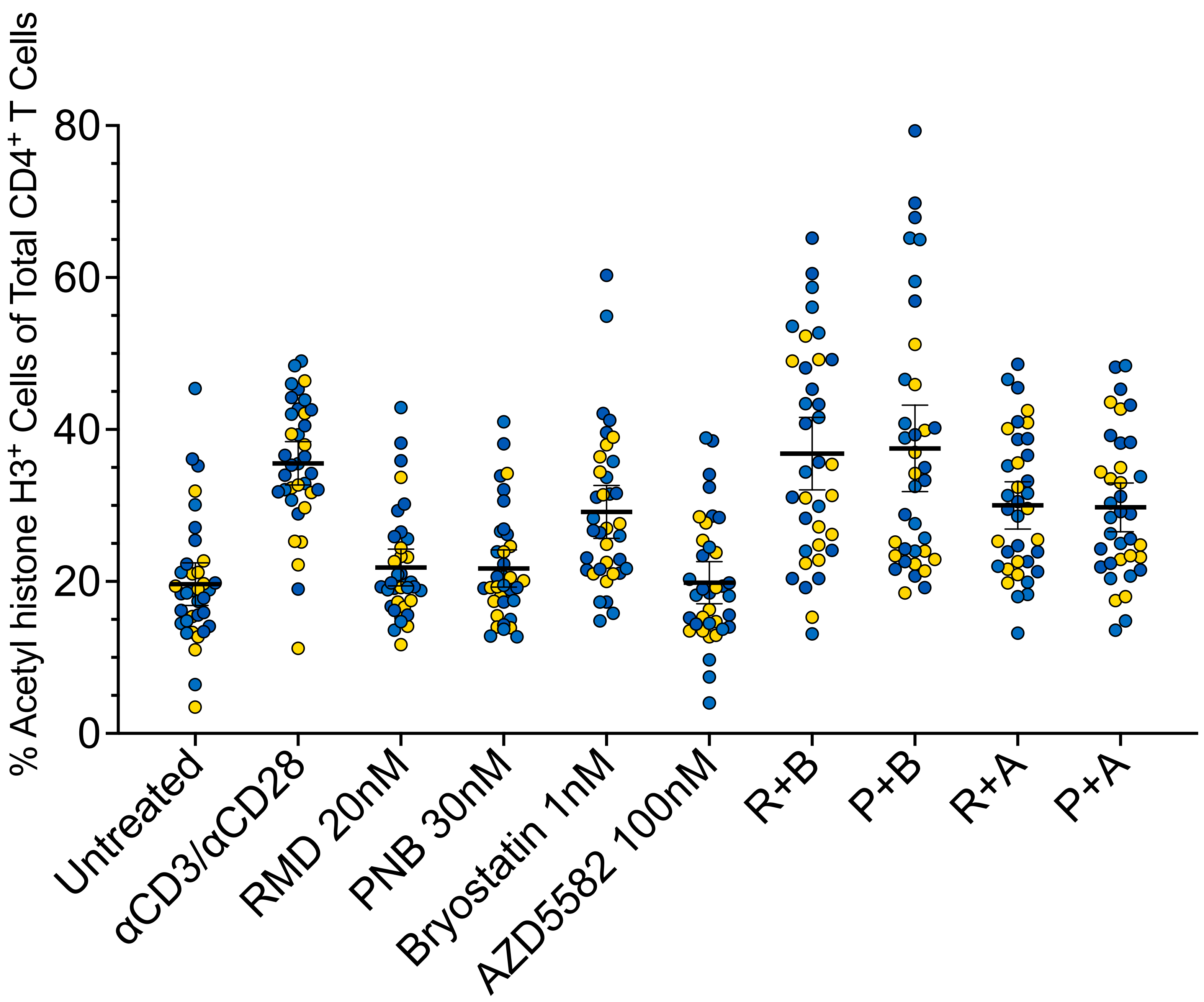


**b.**

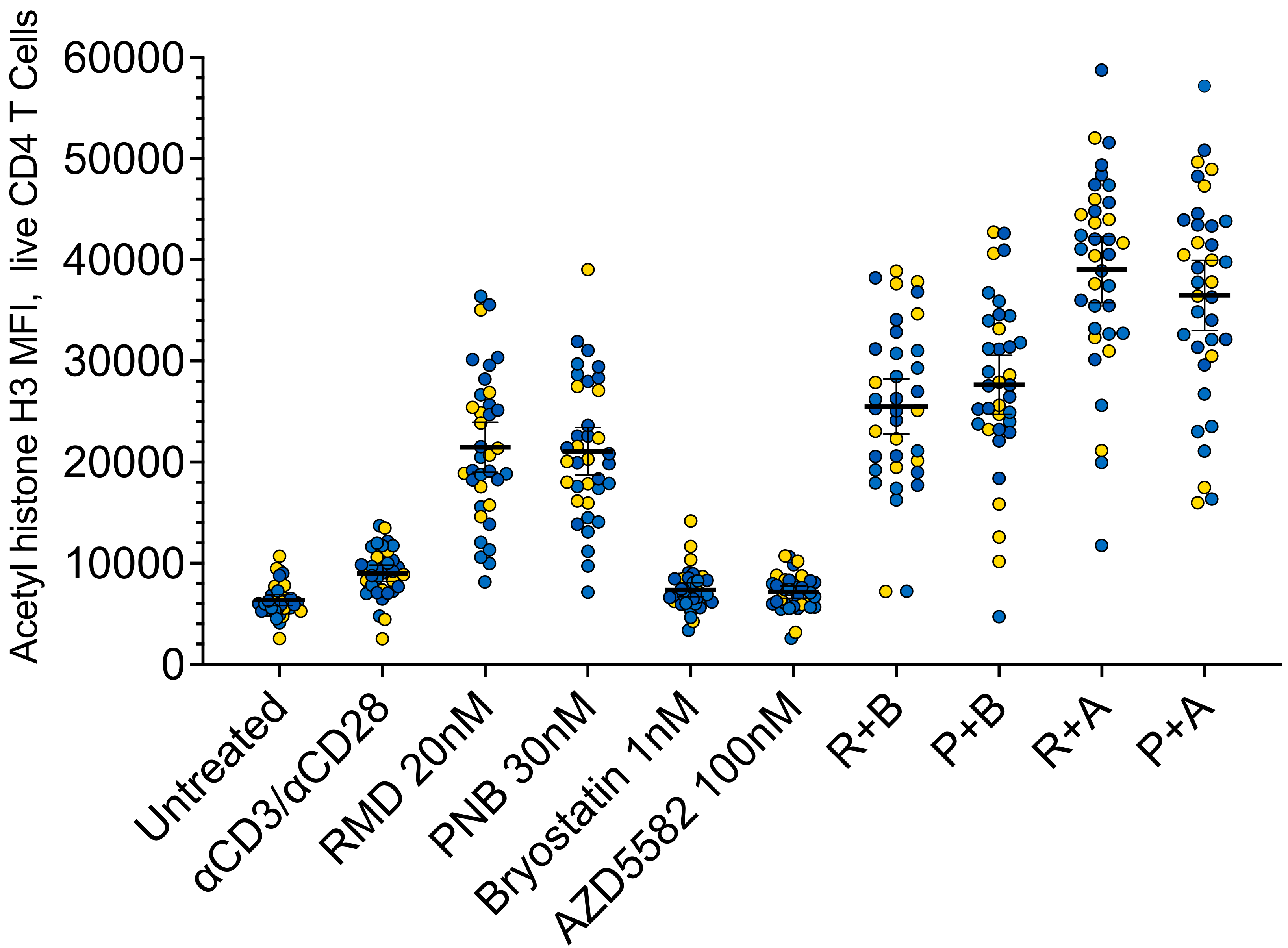

### Supplemental Figure 8

Supplementary Figure 8

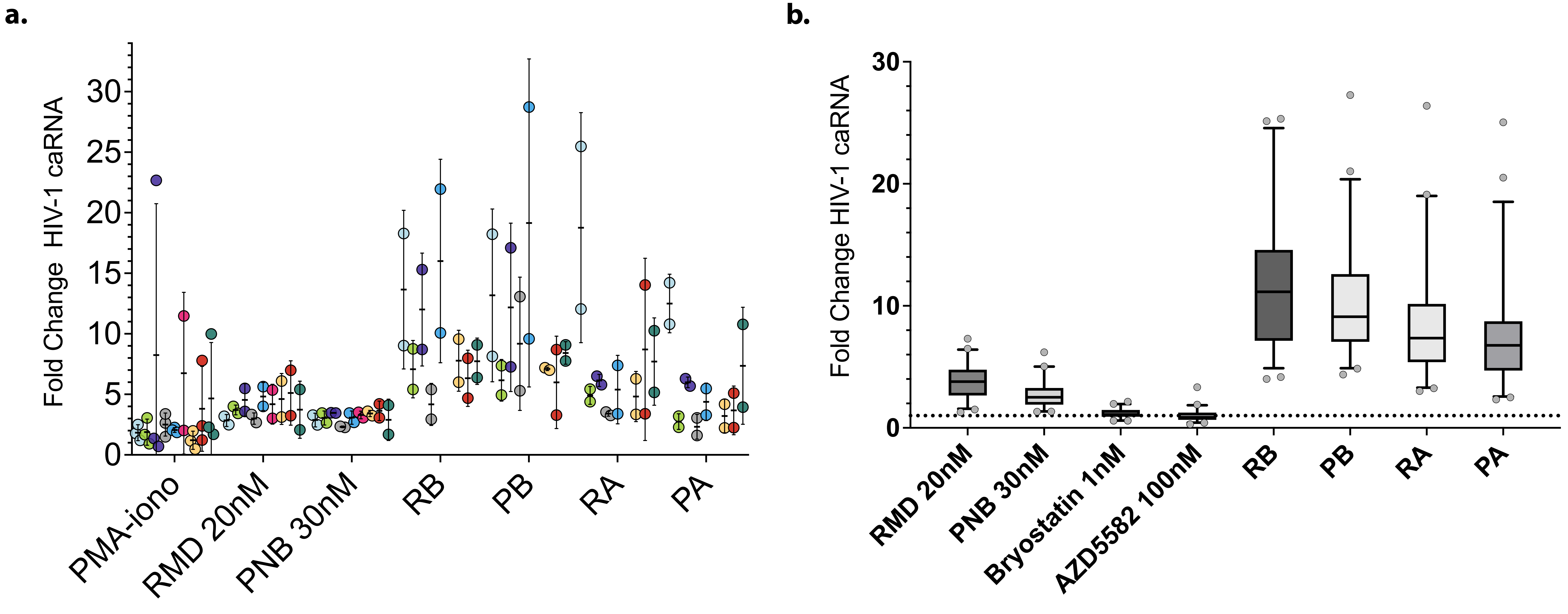
