## Supplemental Figure 7 for "HIV-1 latency reversal agent boosting is not limited by opioid use"

### Supplementary Figure 7

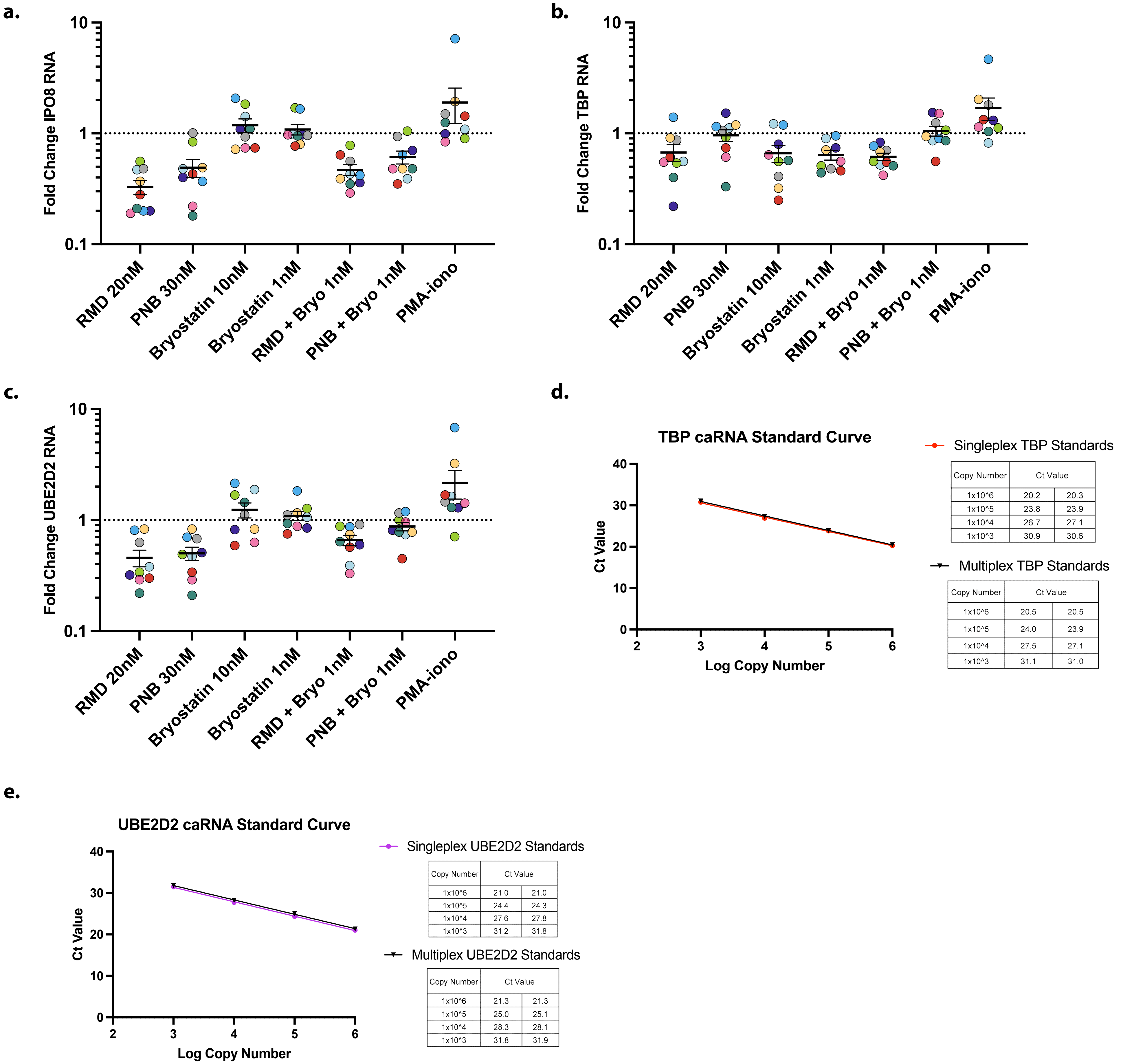


Singleplex TBP Standards

| Copy Number | Ct Value | |
|---|---|---|
| 1x10^6 | 20.2 | 20.3 |
| 1x10^5 | 23.8 | 23.9 |
| 1x10^4 | 26.7 | 27.1 |
| 1x10^3 | 30.9 | 30.6 |

Multiplex TBP Standards

| Copy Number | Ct Value | |
|---|---|---|
| 1x10^6 | 20.5 | 20.5 |
| 1x10^5 | 24.0 | 23.9 |
| 1x10^4 | 27.5 | 27.1 |
| 1x10^3 | 31.1 | 31.0 |

Singleplex UBE2D2 Standards

| Copy Number | Ct Value | |
|---|---|---|
| 1x10^6 | 21.0 | 21.0 |
| 1x10^5 | 24.4 | 24.3 |
| 1x10^4 | 27.6 | 27.8 |
| 1x10^3 | 31.2 | 31.8 |

Multiplex UBE2D2 Standards

| Copy Number | Ct Value | |
|---|---|---|
| 1x10^6 | 21.3 | 21.3 |
| 1x10^5 | 25.0 | 25.1 |
| 1x10^4 | 28.3 | 28.1 |
| 1x10^3 | 31.8 | 31.9 |
