## Supplemental Table 1 for "HIV-1 latency reversal agent boosting is not limited by opioid use"

**Supplementary Table 1. OPHION Opioid Use Sub-Group Characteristics**

| Characteristic | Active injection | Methadone | Suboxone | Chronic Pain | No opioid | **Total*** |
|---|---|---|---|---|---|---|
| Participant, N | 4 | 4 | 12 | 4 | 12 | 36 |
| Age (years) | | | | | | |
| Median (IQR)† | 56 (46-61) | 59 (55-65) | 61 (57-67) | 61 (57-67) | 64 (59-65) | 59 (54-65) |
| Sex | | | | | | |
| Male N (%) | 4 (100%) | 2 (50%) | 9 (75%) | 2 (50%) | 7 (58%) | 24 (67%) |
| Race | | | | | | |
| Black N (%) | 1 (25%) | 1 (25%) | 3 (25%) | 1 (25%) | 9 (75%) | 15 (42%) |
| White N (%) | 1 (25%) | 2 (50%) | 6 (50%) | 2 (50%) | 2 (17%) | 11 (31%) |
| Hispanic/Latino N (%) | 0 | 1 (25%) | 1 (8.3%) | 0 | 1 (8%) | 3 (8%) |
| American Indian N (%) | 0 | 0 | 0 | 1 (25%) | 0 | 1 (3%) |
| Declined/NA | 2 (50%) | 2 (50%) | 2 (17%) | 0 | 0 | 6 (17%) |
| Ethnicity | | | | | | |
| Non-Hispanic N (%) | 2 (50%) | 1 (25%) | 7 (58%) | 3 (75%) | 11 (92%) | 24 (67%) |
| Hispanic N (%) | 2 (50%) | 3 (75%) | 5 (42%) | 1 (25%) | 1 (8%) | 12 (33%) |
| ART Duration (months) | | | | | | |
| Median (IQR) | 146 (96-209) | 94 (79-111) | 132 (81-167) | 173 (161-188) | 162 (106-204) | 144 (97-179) |
| Duration of virus suppression (months) | | | | | | |
| Median (IQR) | 76 (51-96) | 62 (35-95) | 79 (64-89) | 77 (41-101) | 70 (38-91) | 75 (44-93) |

*Percentage totals may not add up to 100 due to rounding. †IQR, interquartile range
