## Supplemental Table 1 for "HIV-1 latency reversal agent boosting is not limited by opioid use"

**Supplementary Table 2. HEAL Participant Characteristics**

| Characteristic | Cohort |
|---|---|
| Participants, N | 11 |
| Age | |
| Median (IQR†) | 56 (55-62) |
| Sex | |
| Male (%) | 7 (64%) |
| Race | |
| Black N (%) | 6 (55%) |
| White N (%) | 3 (27%) |
| Hispanic/Latino** N (%) | 0 |
| American Indian N (%) | 0 |
| Other | 2 (18%) |
| Ethnicity | |
| Non-Hispanic N (%) | 9 (82%) |
| Hispanic N (%) | 2 (18%) |
| Duration of viral suppression (months) | |
| Median (IQR) | 66 (26-83) |

†IQR, interquartile range. ** Participants who reported Hispanic ethnicity identified their race as "Other."
